# Can polygenic risk scores contribute to cost-effective cancer screening? A systematic review

**DOI:** 10.1101/2021.11.26.21266911

**Authors:** Padraig Dixon, Edna Keeney, Jenny C. Taylor, Sarah Wordsworth, Richard M. Martin

## Abstract

Polygenic risk is known to influence susceptibility to cancer. The use of data on polygenic risk, in conjunction with other predictors of future disease status, may offer significant potential for preventative care through risk-stratified screening programmes. An important element in the evaluation of screening programmes is their cost-effectiveness.

We undertook a systematic review of papers evaluating the cost-effectiveness of screening interventions informed by polygenic risk scores compared to more conventional screening modalities. We included papers reporting cost-effectiveness outcomes in the English language published as articles or uploaded onto preprint servers with no restriction on date, type of cancer or form of polygenic risk modelled. We excluded papers evaluating screening interventions that did not report cost-effectiveness outcomes or which had a focus on monogenic risk. We evaluated studies using the Quality of Health Economic Studies checklist.

Ten studies were included in the review, which investigated three cancers: prostate (n=5), colorectal (n=3) and breast (n=2). All study designs were cost-utility papers implemented as Markov models (n=6) or microsimulations (n=4). Nine of ten papers scored highly (score >75 on a 0-100) scale) when assessed using the Quality of Health Economic Studies checklist. Eight of ten studies concluded that polygenic risk informed cancer screening was likely to be more cost-effective than alternatives. However, the included studies lacked robust external data on the cost of polygenic risk stratification, did not account for how very large volumes of polygenic risk data on individuals would be collected and used, did not consider ancestry-related differences in polygenic risk, and did not fully account for downstream economic sequalae stemming from the use of polygenic risk data in these ways. These topics merit attention in future research on how polygenic risk data might contribute to cost-effective cancer screening.

**Funding:** This work was supported by Cancer Research UK under grant number C18281/A29019. PD and RM are members of the MRC Integrative Epidemiology Unit at the University of Bristol which is supported by the Medical Research Council and the University of Bristol (MC_UU_12013/1, MC_UU_12013/9). PD, EK ad RMM received support from a Cancer Research UK (C18281/A29019) programme grant (the Integrative Cancer Epidemiology Programme). SW and JCT receive funding from the Oxford NIHR Biomedical Research Centre.

**Registration:** Prospectively registered on PROSPERO database before searches commenced. Available at https://www.crd.york.ac.uk/prospero/display_record.php?RecordID=243659

## 1 Introduction

### 1.1 Rationale

This paper systematically reviews the literature assessing the use of polygenic risk data to influence the cost-effectiveness of cancer screening. Genetic testing for inherited cancer susceptibility is an established part of care for individuals where family histories or ancestry indicate significant liability to cancer in many health care systems [1]. For example, genetic data can be used to inform the future risk of disease as part of cancer screening programmes in populations with founder mutations, such as screening interventions directed at identifying carriers of BRCA1/BRCA2 breast and ovarian cancer susceptibility mutations amongst Ashkenazi Jews [2].

However, it is increasingly appreciated that polygenic as well as monogenic risk may affect cancer incidence and its progression [3]. Polygenic risk reflets the cumulative influence of many different sources of genetic variation on disease risk, rather than the influence only of rare pathogenic mutations in single genes such as, for example, the BRCA1/BRCA2 genes for breast and ovarian cancer [4].

Polygenic risk scores (PRSs) reflect the aggregated effect of many genetic variants (typically single nucleotide polymorphisms) that are known to influence the incidence of cancer or some other cognate outcome [5]. PRSs may be predictive of disease risk, even if the individual variants of which it is comprised themselves have only modest impacts on risk when considered alone [6-8].

Across many individuals, a cancer PRS will exhibit a distribution of risk that reflects higher and lower genetic susceptibility to the specific cancer analyzed. For example, Callender et al [9] reported that individuals in the first and 99^th^ percentiles for PRS of incident prostate cancer have relative risks of 0.09 and 5.52 respectively, compared to population means. In some cases, differences in relative risk are associated with potentially significant differences in absolute risk. For example, Mavaddat et al [10] found that the estimated lifetime absolute risk of ER-positive breast cancer by age 80 years ranged between 2% for women in the lowest centile of polygenic risk to 31% of those in the highest centile.

Polygenic risk appears in some cases to influence the risk associated with monogenic risk variants [11], and maybe used to stratify risk and refine treatment decisions for individuals with monogenic mutations [12]. Kapoor et al [13] found that preventive strategies intended to modify individual risk factor for breast cancer could have a greater influence on absolute risk of breast cancer for women at higher polygenic risk. There is also some evidence that PRS data can help to identify more aggressive tumours [5]. Amongst other uses of these data, they may have utility in screening programmes to identify asymptomatic individuals at increased risk of incident disease [14-17]. This could involve better identification of those at the highest absolute risk for incident cancer, the prediction of treatment response using PRSs, or other means of improving the expected benefits of screening.

Conversely, there are several reasons why polygenic risk data might not necessarily improve the effectiveness and cost-effectiveness of screening programmes. Most individuals will not be in the tails of a polygenic risk distribution. Any modulation of baseline absolute cancer risk that can be obtained from improved knowledge of polygenic risk may be limited across a population, since even large inter-percentile relative risks may be associated with modest differences in absolute risk [17, 18]. These issues are compounded by the modest heritability of many cancers, false positives associated with polygenic risk profiling. There are circumstances where polygenic risk offers little incremental enhancement to prediction of risk amongst those carrying penetrant monogenic mutations, a positive family history, or other readily available risk factors [19].

These considerations have an important bearing on the potential uses of polygenic risk data in cancer screening. Screening generally requires clear evidence of a net benefit to the wider community to justify invasive and potentially harmful investigations such as biopsy and the possibility of overtreatment of indolent cancers [20]. These and other considerations reflect the established Wilson-Jungner criteria [21] for screening in both cancer and non-neoplastic disease, which recommend that screening programmes be cost-effective and are therefore efficient uses of scarce healthcare resources that may have more valuable alternative uses [22]. Similarly, the European Guide on Quality Improvement in Comprehensive Cancer Control [23] recommends that the cost-effectiveness of screening interventions be evaluated before intervention implementation.

The use of polygenic risk may influence the cost-effectiveness of cancer screening in a number of ways, but will likely stem from targeting resources at individuals with the greatest need of intervention. For example, Callender [9] found that there is 50% less over-diagnosis of prostate cancer of men in the highest quartile of polygenic risk for this cancer compared to those in the lowest risk quartile.

However, there could also be many challenges of using PRS routinely in health care. For example, obtaining polygenic risk data (especially on a large scale) be expensive, may lead to higher rates of false positives, over-treatment, increased patient anxiety and higher caseloads for medical professionals without clinical benefit. Moreover, population-level screening using PRSs is likely to be more complex in admixed societies since the great majority of existing PRSs are more predictive of disease in individuals of European ancestry [24].

The fundamental question in screening cost-effectiveness is whether the potential incremental improvement from polygenic risk data improves patient outcomes sufficiently relative to the costs and consequences of acquiring and using these data. A recent review [25] of the use of PRSs in screening found only very limited investigation of the cost-effectiveness of using PRSs in screening, and no systematic reviews of the use of PRS in cancer screening have been undertaken. The 2021 Polygenic Risk Score Task Force of the International Common Disease Alliance [5] noted the very limited economic evidence concerning uses of PRSs in general and recommended that research address this as a priority using studies that “consider both economic factors and factors and healthcare management that vary across clinical settings and region”.

To address some of this evidence gap, this paper reports our systematic literature review which assessed whether using polygenic risk data is likely to influence the cost-effectiveness of cancer screening.

### 1.2 Objectives

Our objectives were to assess the extent of the literature evaluating the use of PRSs in cost-effectiveness analyses of cancer screening, to examine how PRSs were used in cost-effectiveness cancer screening models, and to evaluate how PRS data influences the cost-effectiveness of cancer screening interventions compared to non-cancer screening modalities. Screening is a process to identify individuals, who may be asymptomatic, at increased risk of disease.

We examined studies with participants undergoing population-scale screening for any form of cancer. Interventions in the scope of our review were cancer screening using PRSs compared to cancer screening without PRSs. We examined outcomes relating to cost-effectiveness measured in terms of incremental cost-effectiveness ratio, cost-benefit ratio, net monetary benefit, net health benefit or other similar statistic. We considered only study designs reporting these types of cost-effectiveness outcome to ensure that we identified quantitative economic evaluations. No restriction was placed on whether cost-effectiveness analyses were informed by or conducted alongside a randomized controlled trial, cohort study, or other study design.

## 2 Methods

We used the PRISMA 2020 systematic review guidelines [26] to inform the development, conduct and reporting of this study. Appendix 1 contains a completed PRISMA checklist.

### 2.1 Protocol and registration

The study protocol is available on the PROSPERO database (https://www.crd.york.ac.uk/prospero/display_record.php?RecordID=243659) to which it was prospectively uploaded in on 1 April 2021 before the first searches commenced on 16 April 2021.

### 2.2 Eligibility criteria

We included studies using PRS data in cost-effectiveness analyses of cancer screening. We considered English-language journal articles or preprints published in any country or time period. Table 1 summarises eligibility criteria.

**Table 1.** Summary of inclusion and exclusion criteria.

| Inclusion criteria (papers included if they meet all criteria) | Exclusion criteria (papers excluded if any of these criteria are encountered) |
| --- | --- |
| **Participants:** Studies including participants undergoing screening for any form of cancer | **Participants:** Participants screened for diseases other than cancer; non-human studies |
| **Intervention & Comparison:** Use of PRS in cancer screening cost-effectiveness evaluation compared to any other non-PRS screening modality | **Intervention & Comparison:** Use of monogenic rather than polygenic tests; no use of genetic tests Interventions not involving screening |
| **Outcomes:** Cost-effectiveness outcome statistics and other outcomes detailed in Section 2.2 | **Outcomes:** Studies not reporting cost-effectiveness outcomes |
| **Study design:** Cost-effectiveness analysis reporting an outcome such as an incremental cost-effectiveness ratio, net monetary benefit, net health benefit. Trial or model-based evaluation using data from any study design | **Study design:** Studies other than cost-effectiveness analyses; cost-effectiveness work not reporting comparisons of incremental measures of cost and effect. |
| **Other criteria:** Any country, health system, or time period; Journal articles; preprints, English language | **Other criteria:** Grey literature. Books. Not in English language. Expert opinion, abstracts, conference proceedings, methodological, general, commentary and review articles not containing original research. |

### 2.3 Information sources

A systematic review of the literature using the NHS Economic Evaluation Database, Medline, EMBASE, HTA databases, NICE guidelines, UK National Screening Committee guidance, preprint servers Bioarxiv and Medrxiv, and hand searches was carried out, with no restriction on date. Relevant data were extracted and the results narratively synthesized.

### 2.4 Search strategies

The full search strategies for all databases are presented in Appendix 2.

### 2.5 Study selection

One author (PD) independently selected reports that appeared to fulfil inclusion criteria based on a review of abstracts and titles. Papers were retained for full-text review when they appeared to meet inclusion criteria or where there was insufficient evidence to exclude them. Two reviewers (PD and EK) then reviewed the full text of articles. Following discussion and reconciliation of any discrepancies in judgments reached by the two reviewers, the full text of articles meeting these criteria was then subject to further review by both reviewers against all inclusion criteria.

### 2.6 Data collection process

Data extraction was informed by the recommendations and example data extraction form proposed by the Centre for Reviews and Dissemination at the University of York [27]. We piloted an initial Microsoft Excel-based data extraction form on a specimen cost-effectiveness analysis. We then refined this based on discussions between two authors (PD and EK). Data were independently extracted to populate this form by PD and EK.

### 2.7 Data items

We obtained the following information from papers meeting all inclusion criteria.

– Study objective
– Cancer(s) studied
– Context (screening strategies compared and country)
– Type of economic evaluation used
– Proposed design for a screening programme (for example is population-level screening of all adults assumed?)
– Risk thresholds (if used)
– Adherence to screening
– Screening interval modelled
– Structure of the economic model
– Age range of cohort
– Size of cohort modelled
– Perspective of the analysis
– Cancer treatments modelled
– Modelling of cancer progression
– Mortality measures considered
– Health state utility values considered
– Duration of follow-up modelled
– Outcome measure
– How genetic data were obtained (or assumed to be obtained) and analyzed
– Assumptions made in creating the polygenic risk score
– Cost of the PRS and any associated costs
– How PRS data were included and modelled
– Whether ethnicity was considered in relation to PRS, and whether differential cost-effectiveness was considered by ethnicity
– Cost-effectiveness threshold used
– Cost-effectiveness results of PRS-informed screening compared to non-PRS screening modalities
– Sensitivity of cost-effectiveness results to model parameters.

### 2.8 Risk of bias and quality assessment in individual studies

We appraised the quality of included studies using the Quality of Health Economics Studies (QHES) checklist [28]. This is not a tool for assessing risk of bias *per se*, but was found to have the highest construct validity amongst 79 tools for assessing the quality of cost-effectiveness studies in a recent systematic review [29].

The QHES uses a weighted grading system to measure the quality of studies against 16 criteria, each of which is weighted by importance. The full list of criteria and their weights is presented in Appendix 3. The scoring system of this checklist is binary: studies receive either the full weighted criterion score or zero depending on whether the particular criterion is met. These criteria include domains relating to study characteristics, study design, and results and conclusions. The range of the QHES score is from 0 to 100; values over 75 indicate a study of high quality. Each study was assessed using the QHES checklist by one author (PD).

### 2.9 Summary measures

We extracted data on the cost-effectiveness of cancer screening interventions using PRSs in comparison to interventions without PRS data. This included data on incremental cost-effectiveness ratios, net benefit, and related summary measures of intervention cost-effectiveness. We also extracted and summarised information on the methods by which PRS data were used in each cost-effectiveness model implementation as detailed in Section 2.7 above.

### 2.10 Synthesis of results and additional analyses

A narrative synthesis of results was undertaken. No additional analysis such as subgroup analysis or meta-regression was planned or undertaken. Where necessary, we referred to literature cited in included to clarify any missing or unclear data issues. Certainty assessment involved consideration of the limitations of the included literature.

## 3 Results

### 3.1 Study selection

Searches concluded in June 2021 and reflect literature that met the inclusion criteria up to that point. The systematic search, undertaken using the data-specific search terms set out in Appendix 2, identified 660 articles (Figure 1, based on the PRISMA 2020 statement [26]).

**Figure 1.**
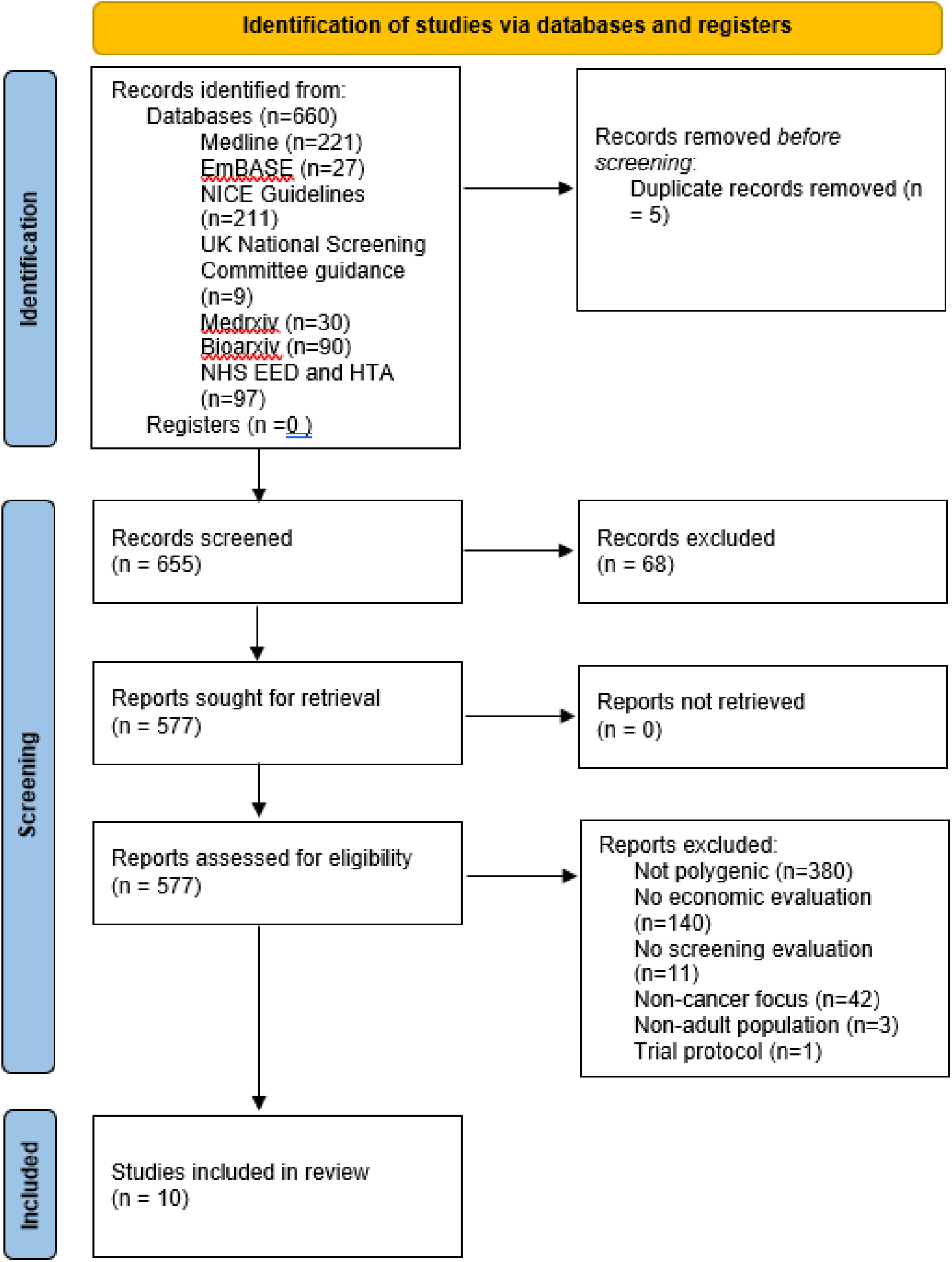
Identification of studies.

Excluding duplicates, the titles and abstracts of 665 articles were screened, from which 68 records were excluded because they were denoted as review articles (n=63), or comprised an abstract only (n=5). After this step, 577 records were successfully sought for retrieval and assessed for eligibility. A total of 567 records were then excluded, the most frequent reasons for which were an absence of polygenic focus or the absence of an economic evaluation. Ten studies that met inclusion criteria were included in the review.

### 3.2 Study characteristics

Table 2 summarises *some* of the key characteristics of each of the ten included papers. A *full* table detailing all study characteristics is presented in Appendix 4.

**Table 2.**
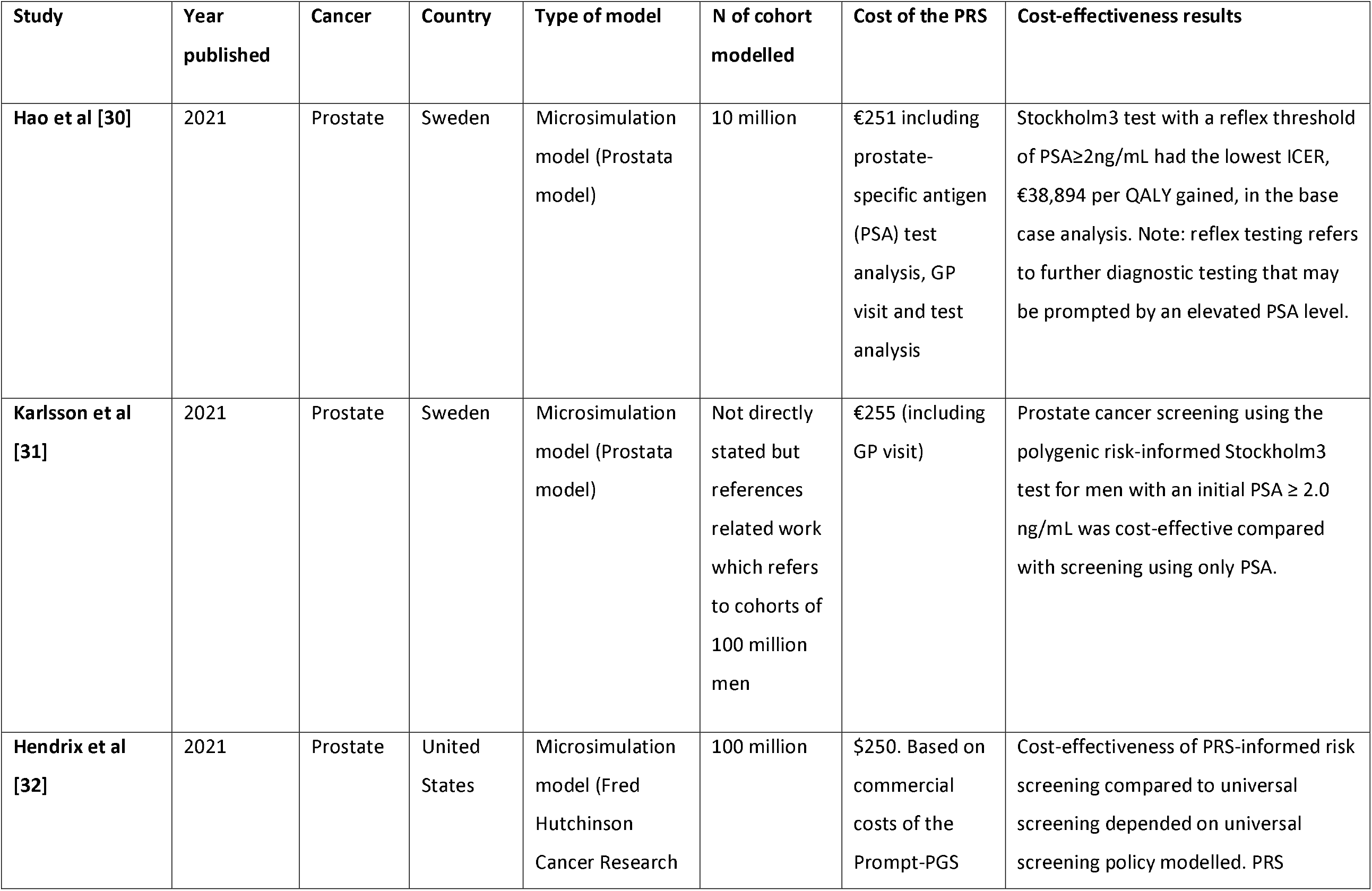
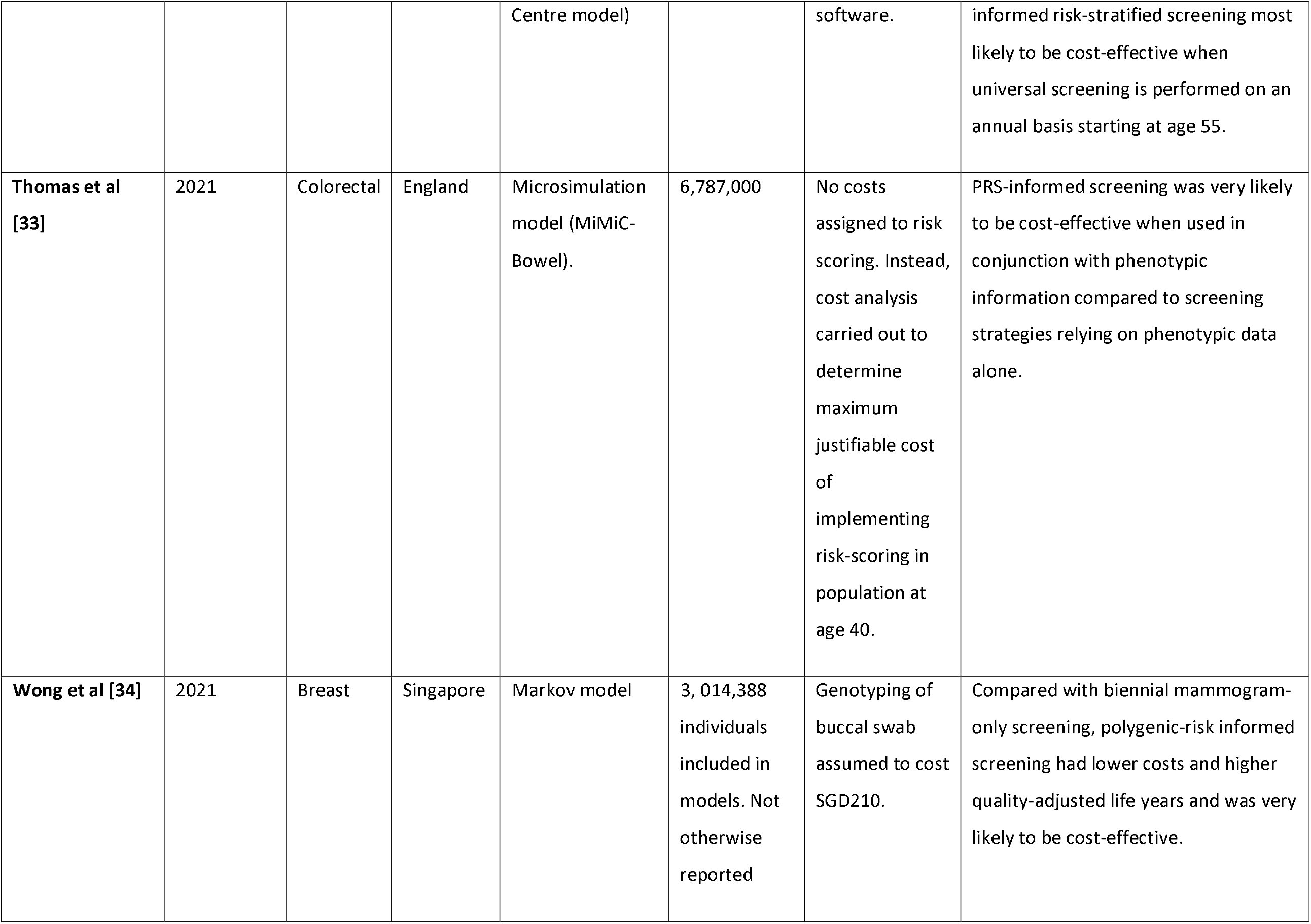
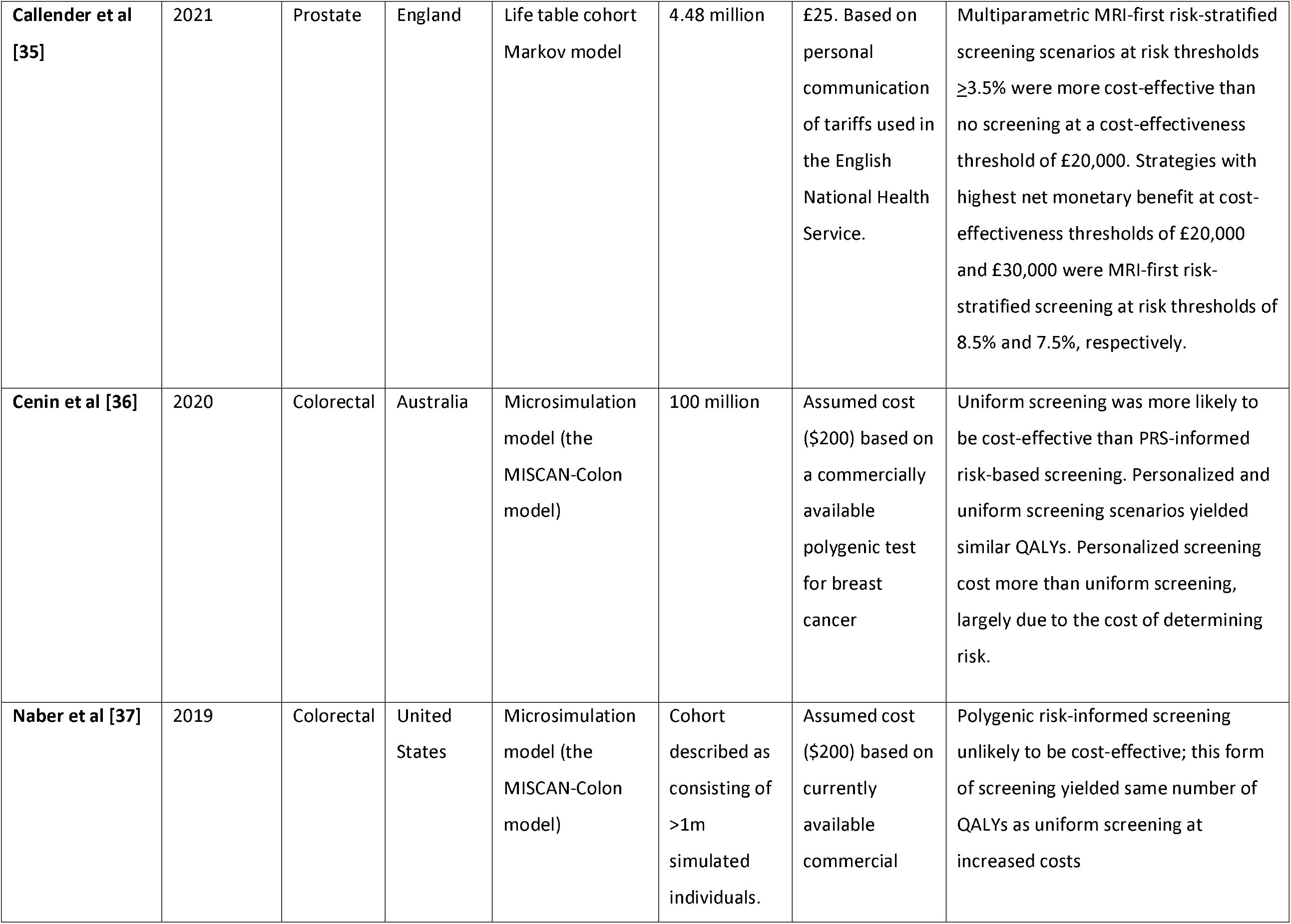
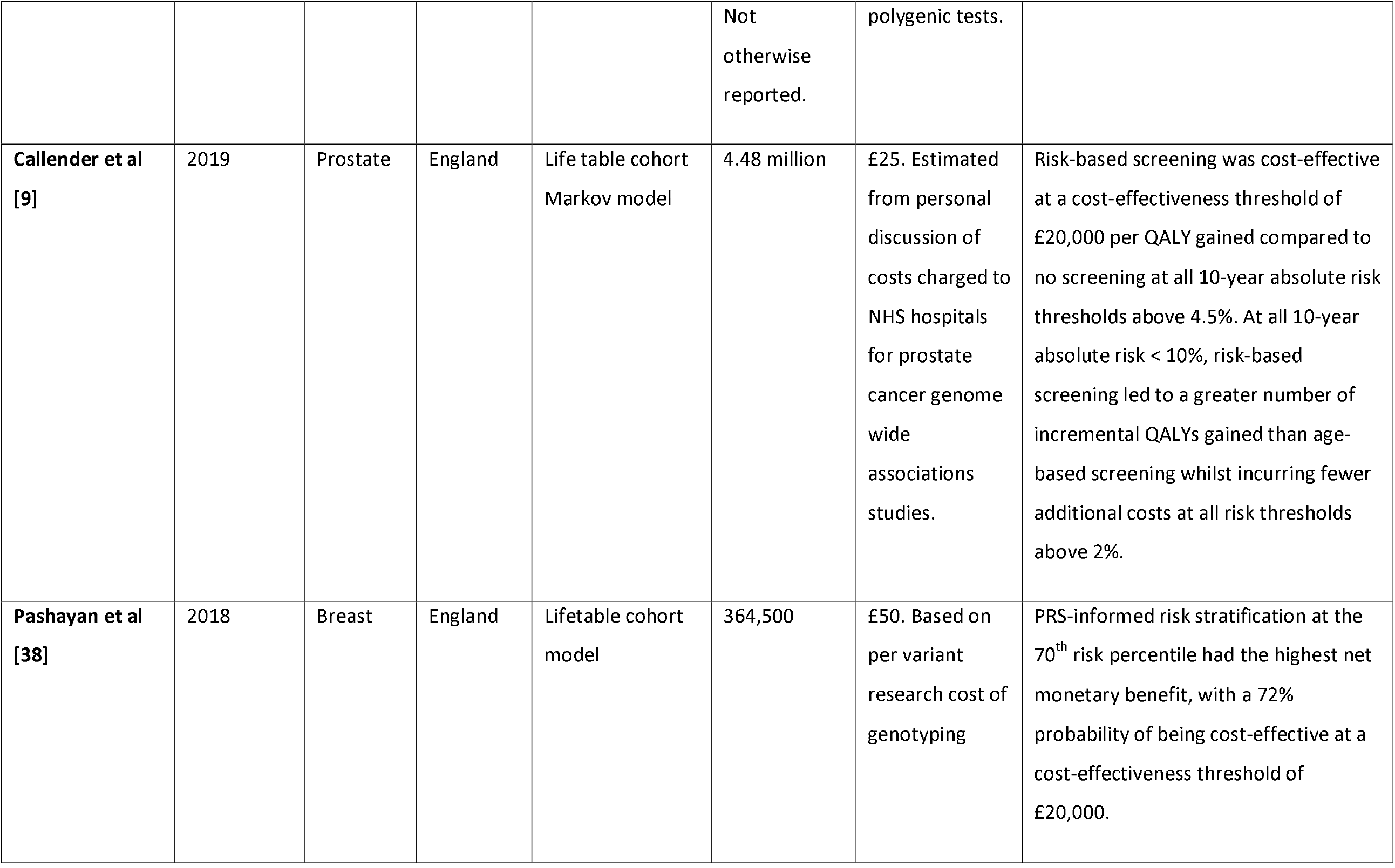
Selected characteristics and outcomes of included studies

Included papers were recently published, with the oldest paper published in 2018. Three cancers were studied in these ten papers: prostate cancer (n=5), colorectal cancer (n=3) and breast cancer (n=2). All studies used a cost-utility design, in which costs were related to quality-adjusted life years (QALYs). This permits the comparison of the cost-effectiveness of these cancer screening programmes with other types of intervention in different clinical areas. Seven studies used a health system perspective to define the scope of costs to be included in these analyses. The exceptions were Naber et al [37] and Hao et al [30] (both health system and societal perspectives) and Karlsson et al [31] (societal perspective).

These cost utility models were implemented using cohort models (n=6), or microsimulation models (n=4). Both cohort models and microsimulation models involve a dynamic simulation of health and disease processes over time. Microsimulation models generally permit greater flexibility in the modelling of event timing and with respect to the interdependency of events [39]. In both types of model, each simulated individual sojourns for a period of time in different states of health, to which are attached state-specific costs and quality of life values. These state-specific values are combined with the amount of person-time spent in each state to produce cohort-level cost-effectiveness parameters, such as incremental cost-effectiveness ratios and net monetary benefit statistics, which enable comparisons of different screening strategies.

Conventional cost-effectiveness thresholds (which are used to inform funding decisions in health technology assessments) were applied in the seven papers that used these thresholds in ex-ante analysis. Wong et al [34] and Naber et al [37] calculated cost-effectiveness thresholds in relation to other model parameters. Hendrix et al [32] compared strategies on the basis of incremental cost-effectiveness ratios and the associated concept of dominance.

Baseline health state utility values were drawn in six papers from population values. The exceptions were Wong et al [34], Hendrix et al [32], Naber et al [37] and Cenin et al [36]. Adjustments were made in all papers for utility in different health states, and for disease progression, interventions and treatments. No adjustments, whether positive or negative, were made to utility as a result of an individual’s knowledge of their own polygenic risk in any of the papers.

In each type of model, cancer treatments were typically defined by the stage of cancer and by applicable local or national guidelines on cancer care. The microsimulation models generally offered more detailed modelling of the natural history of cancer progression. The populations modelled reflected profiles of those typically eligible for inclusion in cancer screening in the specific jurisdiction studied, and followed up simulated individuals for an appropriate amount of time, including lifetime follow-ups. The screening intervals modelled broadly reflected “real world” screening implementations. This is important since the choice of comparator intervention has a direct and material impact on the estimated cost-effectiveness of an intervention in any comparative incremental analysis.

None of the papers described in any detail how genetic data for the entire modelled cohort would be acquired. There may be significant impacts on cost-effectiveness depending on when and where in the clinical pathway the sample to create the PRS would be obtained, which genotypes would be assessed, how the results would be interpreted and how findings would be communicated. Some papers ([30, 31, 35]) noted that a primary care consultation as a first step in the process.

There was no modelling of the impact of differential polygenic risk by ancestry. Six papers modelled the effect of lower take-up or imperfect adherence to screening; the exceptions were Wong et al [34], Hao et al [30], Karlsson et al [31] and Hendrix et al [32].

Various means of including polygenic risk and assessing their influence in relation to risk thresholds or other indicators for intervention were implemented. Wong et al [34] created tertiles of a hypothetical polygenic risk distribution to identify low, medium and high polygenic risk groups. Hendrix et al [32] evaluated the impact of the proprietary Prompt Prostate Genetic Score (PGS)®. The number of alleles is not described, but references in that paper suggest that it is based on 29 prostate cancer SNPs in 4,528 men of European ancestry the placebo arm of the Prostate Cancer Prevention Trial [40].

Hao et al [30] and Karlsson et al [31] evaluated the Stockholm3 package. This model combines measurement of PSA, protein biomarkers, polygenic risk scores (based on 232 SNPs) and clinical information collected by questionnaire, including age, family history and previous prostate biopsies, [41]. These variables have all been reported to be associated with prostate cancers with a Gleason score >7.0 and the Stockholm3 model has been shown to outperform PSA testing alone in predicting prostate cancers with Gleason>7 [41]. Karlsson et al [31] evaluated screening strategies that used Stockholm3 test as a reflex test (i.e. tests prompted after particular levels of PSA) for PSA values ≥ 1, 1.5 and 2 ng/mL. Hao et al [30] implemented a similar model with a screening strategy that used Stockholm3 with reflex test thresholds of PSA≥1.5 and 2ng/mL

Naber et al [37] generated a relative risk distribution with different values of the area under the curve (AUC) for a polygenic test ranging from 0.60 to 0.80 in each simulated, hypothetical cohort. These cohorts were split in 60 groups defined by their relative risk of colorectal cancer.

Thomas et al [33] modelled random assignment of risk alleles to individuals to reflect allele frequencies obtained from UK Biobank data, as well as correlations between alleles on the same chromosome. Genetic [42] and non -genetic risk factors were combined to obtain individualised relative risks for colorectal cancer, which were applied to transition probabilities from normal epithelium to adenoma and to colorectal cancer. Estimated relative risk was adjusted to ensure that the simulated distribution of disease reflected expected colorectal cancer incidence.

Cenin et al [36] stratified their cohort population into five risk groups based on quintiles of polygenic risk and fist degree family history of colorectal cancer. The prevalence of the five categories was simulated given a 20% probability of being in any SNP quintile and a 10% probability of positive first-degree family history. The relative risk of colorectal cancer, compared to average population risk, was based on combined relative risk of each polygenic risk quintile and family history.

Pashayan et al [38], Callender et al [9] and Callender et al [35] followed a similar approach. Given a log-additive model of interaction between genetic and conventional non-genetic risk factors, the distribution of risk for incident disease is log-normal on a relative risk scale. Callender et al [9] used data from Schumacher et al [43] and Dadaev et al [44]. Percentile ranks associated with relative risk or absolute risk were obtained given knowledge of the mean and variance of the log-normal relative risk distribution. This permitted, for example, estimation of the proportion of a population above or below a particular risk threshold.

### 3.3 Risk of bias/study quality within studies

We evaluated all studies using the Quality of Health Economics Studies (QHES) checklist [28]. Appendix 5 in the supplementary material details the score awarded to each study against the 16 criteria of this checklist. The QHES checklist does not identify the presence or scale of biases that may affect the conclusions of the included studies. However, it does permit a characterisation of the quality of the economic evaluation and the implications that this may have for the robustness of results, allowing for some degree of ineradicable subjectivity in assessing studies against each quality criterion.

All but one study (Wong et al [34]) received a score indicating high study quality (>75 on a 0-100 scale). Wong et al [34]) did not reach the threshold for a number of reasons, but included a the absence of remission from cancer and of the possibility of between-state transitions in their model. We concluded that even the highest scoring studies did not account for all potential downstream impacts of PRS-related interventions. This is an inherently difficult task but its identification does indicate the qualifications that must be used in interpreting each paper and suggests one possible area on which future research might focus.

### 3.4 Results of individual studies and synthesis of results

We extracted summary information on the cost-effectiveness of polygenic risk-informed screening strategies compared to strategies that did not use these data. It was not feasible to explore heterogeneity in the cost-effectiveness conclusions across studies given the relatively small number of papers meeting inclusion criteria and given the narrow range of cancers studied. Direct comparison across studies is also complicated by the differences in how cost-effectiveness results were reported, by differences in the ways in which polygenic risk was incorporated into wider risk models, and in types of strategy compared.

Eight of the ten studies concluded that polygenic risk-informed screening was likely to be cost effective, or had the lowest incremental cost-effectiveness ratio of strategies evaluated. The two exceptions were Naber et al [37] and Cenin et al [36].

Conclusions in all ten included papers were conditional on important model parameters such as the cancer studied, risk thresholds, screening uptake and compliance, assumptions regarding background testing of polygenic risk outside screening programmes, comparator interventions, age at which screening commences, screening intervals and other model parameters. Cost-effectiveness results in all papers other than Wong et al were reported to be somewhat sensitive to the cost of polygenic risk stratification. Where assessed, results were also sensitive to the discrimination of polygenic risk stratification. Note that these sensitivity analyses of model parameters were deterministic or “one-way”. This approach can be misleading given correlations between parameters and the non-linear structure of the various simulation models used [45].

## 4 Discussion

### 4.1 Summary of evidence

We conducted and reported the first systematic review of the cost-effectiveness of using polygenic risk data in population-scale cancer screening in comparison to more conventional screening modalities. The evidence base in this area is both recent and relatively small. We identified ten studies encompassing three different cancers that met our inclusion criteria. Most studies concluded that the use of polygenic risk to inform risk stratification for cancer screening was likely to be cost-effective.

Each study used varieties of simulation models (either cohort Markov models or microsimulation models) to capture the dynamic process of disease over time in cost-utility frameworks. Nine of the ten studies were judged to be of high quality when assessed against the Quality of Health Economic Studies (QHES) checklist, although these and similar checklists do not capture all considerations relevant to the implementation of PRSs in cancer screening.

### 4.2 Limitations

#### Limitations of the evidence

The most robust source of evidence on the long-term impact on cost-effectiveness of using polygenic risk data in cancer screening would come from trials with very long follow-up of mortality and other outcomes. In the absence of data from such trials, cost-utility simulation models of the type meeting the inclusion criteria of this review are likely to constitute the best available alternative means of evaluating how PRSs might contribute to cost-effective cancer screening.

Important limitations of the available evidence including open questions on the downstream economic impacts of using PRS, how PRS data might be acquired and used on a large scale and how this might affect the design and implementation of screening programmes, different ways of modelling polygenic risk and its relation to cancer incidence and progression, and structural and parameter uncertainty generally.

##### DOWNSTREAM CONSEQUENCES

It is evident from the review that there is a need for greater evidence on the economic consequences of using PRS in screening, including their acquisition costs and downstream economic sequelae, and remuneration for their integration into routine care. All studies subsumed the economic consequences of polygenic risk-informed screening into a limited number of disease-specific states.

These states did These states did not include potential service re-designs that may be necessary to give implement this type of screening, nor did they specify the service re-designs that may need to be implemented as a consequence of this screening. These could include changes in rates of consultation under more widespread use of polygenic risk data, the cost implications of an education programme for primary and secondary care providers, potential optimizing of prescribing behaviour using pharmacogenomics, and others.

##### COSTS OF PRS

All studies necessarily lacked robust external data on the per-individual costs of polygenic risk stratification for use in large-scale screening programmes. Studies either assumed a cost for obtaining the information necessary to undertake polygenic risk-informed screening, or back-calculated the costs at which such screening might alter estimated cost-effectiveness. Most papers (other than Hendrix et al [32] and Naber et al [37]) parameterised uncertainty around the mean costs of polygenic risk stratification or used a threshold analysis (as in Thomas et al [33] and Cenin et al [36]) to examine the sensitivity of cost-effectiveness results to its cost. All papers reported versions of probabilistic sensitivity analysis but consideration of the specific impact of input parameters such as the cost of risk stratification was generally deterministic, which will tend to understate the degree of uncertainty applying to the model’s conclusions.

##### ACQUIRING AND USING GENETIC DATA ON A LARGE SCALE

None of the papers described in any detail how genetic data for the entire modelled cohort would be acquired. All studies assumed the availability of polygenic risk data with complete coverage of their target population. There was some mention of samples being taken by buccal swab (e.g. Wong et al [34]) or that any test to collect these data would be administered by a general practitioner (e.g. Cenin et al [36]) but there was little to no other consideration given to how these data would be collected at scale.

For example, papers assuming the availability of a comprehensive PRS for middle aged men in England is typically based on genetic analysis conducted on individuals of European ancestry, whereas approximately 7-8% of men in the target age groups did not report this ancestry in the 2011 UK census [46]. This is tantamount to conditioning the analysis on the most predictive PRSs available, even if all members of a simulated cohort would not have had access to such a PRS. More generally, all papers condition on the availability of polygenic risk data at scale – this is not necessarily a criticism of the ten papers meeting our inclusion criteria, but it does limit the direct relevance of this literature to policy decisions concerning the collection and use of population-scale polygenic risk data for cancer screening.

##### STRUCTURAL AND PARAMETER UNCERTAINTY

There little to no assessment of the sensitivity of cost-effectiveness to different ways of modelling PRSs, and the sensitivity of model parameters to PRSs. For example, recent work on breast cancer has indicated that polygenic risk might alter treatment decisions for those individuals with monogenic variants close to thresholds for intervention [12]. More generally, the included papers rely on models that have a primary and often exclusive focus on disease incidence, in which PRS only affects the risk of cancer, with no impact on progression and, in some cases, little or no differentiation of the impact of polygenic risk by grade of cancer.

These forms of long-term economic simulation studies necessarily make a number of assumptions to ensure the feasibility and tractability of their analysis. The conclusions of included studies are therefore also conditional on model structure and model parameters, and For example, further scrutiny of adherence and uptake in screening, particularly if there is differential uptake by mode of screening, may address some of the more significant parameter uncertainties. Likewise, changes in quality of life may be differ by screening modality, if, for example, knowledge of high polygenic risk induces anxiety [19].

Structural assumptions in these types of model include the number of alternatives to which interventions are compared, and the number of disease states modelled. Whereas parameter uncertainty may be accounted for to some degree by probabilistic sensitivity analysis [22, 47], uncertainty about model structure was (at best) approached by investigating results under different assumptions, but without necessarily characterising the plausibility of the scenarios modelled under these assumptions.

#### Limitations of the systematic review

This review assessed literature published or uploaded to preprint servers in the English language by June 2021. Only three cancers were studied in the ten papers included in the review, and it was not possible to infer whether cancer type influenced the probability of polygenic risk-informed screening being cost-effective. Research is moving quickly in this area, and new findings and methodologies are likely to soon emerge. In particular, the identification of new variants influencing disease and the development of new methods to estimate polygenic risk scores may also alter the balance of evidence in this area, particularly if new scores are more predictive.

As the literature develops and the number of papers meeting inclusion criteria grows, it may be feasible to examine potential modifiers of screening cost effectiveness. These may include the type of cancer studied, the broad structure of the screening interventions modelled, the health system in which the screening is to be performed, and the age structure of the population modelled. For example, it is plausible that polygenic influences on disease incidence and progression will differ by cancer, even given further progress in elucidating the polygenic basis of different cancers more generally.

We did not have information on the scale and scope of any publication bias in this area. We did not plan to conduct meta-analysis given anticipated heterogeneity in cancer type, model structure, and the basis by which polygenic risk was modelled. Instead, we have provided a high-level summary of the cost-effectiveness of polygenic risk-informed modalities compared to more conventional screening strategies, but it was not feasible to summarise all possible combinations of polygenic-risk informed screening strategies relative to more conventional alternatives.

Finally, although we used the QHES checklist, which was assessed as having the highest sensitivity of any such checklist included in the systematic review and assessment of Walker et al [29], it does not and cannot identify all issues that may affect the quality and robustness of any particular study. For example, it does not account for all the specific and general limitations identified above, although it does confirm that nine of the ten studies undertook, at a minimum, the fundamental steps needed to undertake a robust economic evaluation.

#### Implications for future research

The systematic review suggests several areas for future research. There is scope to expand the number of cancers studied in this literature, and to extend and improve the sources used to population the parameters of these models to address parameter uncertainty. Future models should also consider how more detailed modelling of natural histories, new screening modalities and diagnostic pathways may influence structural uncertainty although this will likely require some contextual nuance since increasing the complexity of a model does not necessarily secure its robustness or relevance. Recent methodological developments (e.g. [48]) support the use of probabilistic sensitivity analysis, and could be explored in future research to examine sensitivity to key parameters such as the cost of obtaining and using data on polygenic risk.

There is a need to understand the implications of different ways of modelling the influence of polygenic risk, such as going beyond a focus on disease incidence to consider potential impacts of polygenic risk on progression. It is also important to note that different methods for calculating polygenic risk scores at the level of the individual, which will be necessary for risk stratification, may not be stable across methods (in the sense that an individual’s percentile rank may change according to the approach used) and will be associated with uncertainty. Ding et al [49] found large variance in individual PRS estimates, and recommended a probabilistic approach to polygenic risk stratification that estimates the probability that an individual’s PRS is above a pre-specified risk threshold for screening.

There is also a need to establish how the cost-effectiveness of screening might vary by ethnicity given that most available PRSs are most predictive for individuals of European ancestry [24]. Future research may also consider the equity impact of introducing new tests that disproportionately benefits relatively more privileged groups.

One means of addressing the various ways in which cost-effectiveness might be influenced by alternative ways of modelling PRS, improved modelling of ethnicity, and other considerations is the use of comparative modelling as undertaken by, for example, the National Institute’s of Health Cancer Intervention and Surveillance Network [50]. This would involve the similar inputs across a range of models which report the same final outputs. This might facilitate robust comparisons across models that may otherwise differ in their approach to modelling key inputs. Future modelling could also consider the impact of possible intervention-generated inequalities [51] by calculating the required impact of screening interventions in groups (such as those associated with differential coverage of PRSs) that may otherwise face these inequalities.

A further broad area for future research relates to the costing and valuing of polygenic risk data. All studies necessarily lacked robust external data on the per-individual costs of population-level genotyping, and on all downstream economic impacts stemming from the use of these data. Further research is therefore required on the cost of obtaining comprehensive PRS for deployment at the population scale. Health systems generally aim to set remuneration for health technologies at a level that reflects expected population health benefits, expected costs, and system-wide opportunity costs. However, there is an important open question as to whether PRSs are sufficiently different from an economic perspective to other health technologies as to merit distinct treatment by funding and health technology appraisal bodies.

For example, if an individual has their genome sequenced, the marginal cost of producing a PRS for a particular disease is likely to be modest if sufficient economies of scale can be realised in the processing of data already collected. Moreover, the sequenced genetic data is non-rivalrous – unlike conventional economic goods, the use of these data in one context does not prohibit its simultaneous use in another context. These data do not depreciate over time in the way that physical goods may do; indeed, the underlying genetic data may appreciate in value as more associations are uncovered and the uses to which they may be put expand. These features of polygenic risk data challenge conventional approaches to the costing and remuneration of healthcare technologies and merit attention in future research.

### 4.3 Conclusions

The use of polygenic risk data in population-level screening for cancer is attracting increasing interest. A major concern with using these data in population-level screening will be their cost-effectiveness. The literature on this topic is recent, relatively small, and examines only three cancers, albeit that these cancers are relatively prevalent and have screening programmes in some countries. Eight of the ten included studies concluded that the use of polygenic risk data would likely be cost-effective. However, given the limitations identified above, these conclusions should be evaluated in research that addresses limitations and expands the scope of the literature.

Issues that merit particular attention in future research include: modelling of how and where in the care pathway PRSs might be used in screening interventions, understanding how ethnic differences in PRS prediction might affect their use, examining different ways of modelling risk and the impact of structural uncertainty, and ultimately developing further evidence on their wider economic consequences including downstream economic sequelae and integration into very large-scale screening programmes and/or routine care.

This is likely to require prospective evidence from randomized controlled trials (RCTs), as well as comprehensive economic decision-analytic models where several data sources, including RCTs, are synthesised and modelled. Further research is also much needed on how polygenic risk data might be obtained and funded on a population-wide scale, accounting for not only the costs of genotyping or sequencing, but also any related costs such as those associated with changes in rates of healthcare consultations, and interpretation and communication of results to intervention participants.

## Data Availability

The search terms used to interrogate each database are presented in Appendix 2. No other code was used in this review. Template data collection forms are available from the corresponding author on request. All data extracted from each included studied is available in Appendix 3.

## Declarations

The views expressed are those of the authors and not necessarily those of the NHS, the NIHR or the Department of Health or Wellcome Trust.

