## Supplementary material for "Can polygenic risk scores contribute to cost-effective cancer screening? A systematic review"

**Appendix 1 – Completed PRISMA 2020 systematic review checklist**

| **Section and Topic** | **Item #** | **Checklist item** | **Location where item is reported** |
| --- | --- | --- | --- |
| **TITLE** | | |  |
| Title | 1 | Identify the report as a systematic review. | Page 1 |
| **ABSTRACT** | | |  |
| Abstract | 2 | See the PRISMA 2020 for Abstracts checklist. | Page 2 |
| **INTRODUCTION** | | |  |
| Rationale | 3 | Describe the rationale for the review in the context of existing knowledge. | Page 3-6 |
| Objectives | 4 | Provide an explicit statement of the objective(s) or question(s) the review addresses. | Page 6 |
| **METHODS** | | |  |
| Eligibility criteria | 5 | Specify the inclusion and exclusion criteria for the review and how studies were grouped for the syntheses. | Page 6-7 |
| Information sources | 6 | Specify all databases, registers, websites, organisations, reference lists and other sources searched or consulted to identify studies. Specify the date when each source was last searched or consulted. | Page 7-8 |
| Search strategy | 7 | Present the full search strategies for all databases, registers and websites, including any filters and limits used. | Appendix 2 |
| Selection process | 8 | Specify the methods used to decide whether a study met the inclusion criteria of the review, including how many reviewers screened each record and each report retrieved, whether they worked independently, and if applicable, details of automation tools used in the process. | Page 8 |
| Data collection process | 9 | Specify the methods used to collect data from reports, including how many reviewers collected data from each report, whether they worked independently, any processes for obtaining or confirming data from study investigators, and if applicable, details of automation tools used in the process. | Page 8 |
| Data items | 10a | List and define all outcomes for which data were sought. Specify whether all results that were compatible with each outcome domain in each study were sought (e.g. for all measures, time points, analyses), and if not, the methods used to decide which results to collect. | Page 8-9 |
|  | 10b | List and define all other variables for which data were sought (e.g. participant and intervention characteristics, funding sources). Describe any assumptions made about any missing or unclear information. | Page 10 |
| Study risk of bias assessment | 11 | Specify the methods used to assess risk of bias in the included studies, including details of the tool(s) used, how many reviewers assessed each study and whether they worked independently, and if applicable, details of automation tools used in the process. | Page 9-10 |
| Effect measures | 12 | Specify for each outcome the effect measure(s) (e.g. risk ratio, mean difference) used in the synthesis or presentation of results. | Page 10 |
| Synthesis methods | 13a | Describe the processes used to decide which studies were eligible for each synthesis (e.g. tabulating the study intervention characteristics and comparing against the planned groups for each synthesis (item #5)). | Page 10 |
|  | 13b | Describe any methods required to prepare the data for presentation or synthesis, such as handling of missing summary statistics, or data conversions. | Page 10 |
|  | 13c | Describe any methods used to tabulate or visually display results of individual studies and syntheses. | Page 10 |
|  | 13d | Describe any methods used to synthesize results and provide a rationale for the choice(s). If meta-analysis was performed, describe the model(s), method(s) to identify the presence and extent of statistical heterogeneity, and software package(s) used. | Page 10 |
|  | 13e | Describe any methods used to explore possible causes of heterogeneity among study results (e.g. subgroup analysis, meta-regression). | Page 10 |
|  | 13f | Describe any sensitivity analyses conducted to assess robustness of the synthesized results. | Page 10 |
| Reporting bias assessment | 14 | Describe any methods used to assess risk of bias due to missing results in a synthesis (arising from reporting biases). | N/A |
| Certainty assessment | 15 | Describe any methods used to assess certainty (or confidence) in the body of evidence for an outcome. | Page 10 |
| **RESULTS** | | |  |
| Study selection | 16a | Describe the results of the search and selection process, from the number of records identified in the search to the number of studies included in the review, ideally using a flow diagram. | Page 10-12 |
|  | 16b | Cite studies that might appear to meet the inclusion criteria, but which were excluded, and explain why they were excluded. | N/A |
| Study characteristics | 17 | Cite each included study and present its characteristics. | Page 12-15 |
| Risk of bias in studies | 18 | Present assessments of risk of bias for each included study. | Page 18-19 and Appendix 4 |
| Results of individual studies | 19 | For all outcomes, present, for each study: (a) summary statistics for each group (where appropriate) and (b) an effect estimate and its precision (e.g. confidence/credible interval), ideally using structured tables or plots. | Appendix 3 |
| Results of syntheses | 20a | For each synthesis, briefly summarise the characteristics and risk of bias among contributing studies. | Page 16-18 |
|  | 20b | Present results of all statistical syntheses conducted. If meta-analysis was done, present for each the summary estimate and its precision (e.g. confidence/credible interval) and measures of statistical heterogeneity. If comparing groups, describe the direction of the effect. | N/A |
|  | 20c | Present results of all investigations of possible causes of heterogeneity among study results. | N/A |
|  | 20d | Present results of all sensitivity analyses conducted to assess the robustness of the synthesized results. | N/A |
| Reporting biases | 21 | Present assessments of risk of bias due to missing results (arising from reporting biases) for each synthesis assessed. | N/A |
| Certainty of evidence | 22 | Present assessments of certainty (or confidence) in the body of evidence for each outcome assessed. | N/A |
| **DISCUSSION** | | |  |
| Discussion | 23a | Provide a general interpretation of the results in the context of other evidence. | Page 20-23 |
|  | 23b | Discuss any limitations of the evidence included in the review. | Page 20-23 |
|  | 23c | Discuss any limitations of the review processes used. | Page 23-24 |
|  | 23d | Discuss implications of the results for practice, policy, and future research. | Page 24-26 |
| **OTHER INFORMATION** | | |  |
| Registration and protocol | 24a | Provide registration information for the review, including register name and registration number, or state that the review was not registered. | Page 2 |
|  | 24b | Indicate where the review protocol can be accessed, or state that a protocol was not prepared. | Page 2 |
|  | 24c | Describe and explain any amendments to information provided at registration or in the protocol. | N/A |
| Support | 25 | Describe sources of financial or non-financial support for the review, and the role of the funders or sponsors in the review. | Page 2 |
| Competing interests | 26 | Declare any competing interests of review authors. | Page 27 |
| Availability of data, code and other materials | 27 | Report which of the following are publicly available and where they can be found: template data collection forms; data extracted from included studies; data used for all analyses; analytic code; any other materials used in the review. | Page 27 |

**Appendix 2 – Search strategies**

**Medline**

--------------------------------------------------------------------------------

1	cost-effectiveness.mp. or Cost-Benefit Analysis/

2	(cost adj3 (effect$ or util$)).tw.

3	cancer.mp. or *Neoplasms/

4	screening.mp. or *Mass Screening/

5	(Screen$ or test$).tw.

6	Polymorphism, Single Nucleotide/ or Genetic Predisposition to Disease/ or Multifactorial Inheritance/ or Genome-Wide Association Study/

7	1 or 2

8	4 or 5

9	3 and 6 and 7 and 8

**Embase**

--------------------------------------------------------------------------------

1	cost-effectiveness.mp. or exp "cost effectiveness analysis"/

2	(cost adj3 (effect$ or util$)).tw.

3	cancer.mp. or *malignant neoplasm/

4	screening.mp. or cancer screening/ or DNA screening/ or genetic screening/ or screening/ or mass screening/ or screening test/

5	(Screen$ or test$).tw.

6	genetic risk score/ or polygenic.mp. or multifactorial inheritance/ (16396)

7	1 or 2

8	4 or 5

9	3 and 6 and 7 and 8

**CRD HTA,DARE and NHS EED**

Results for: ((cancer) AND (screening ) AND (polygenic OR genetic)) and ((Economic evaluation:ZDT and Bibliographic:ZPS) OR (Economic evaluation:ZDT and Abstract:ZPS) OR Project record:ZDT OR Full publication record:ZDT)

**Medrxiv**

"cancer AND cost-effectiveness AND polygenic"

**Biorxiv**

"cancer AND cost-effectiveness AND polygenic"

**National Institute for Health and Care Excellence (NICE)**

Searched all published NICE guidelines for the word “cancer”

**UK National Screening Committee**

Searched for recommendations pertaining to cancer in adult populations

**Appendix 3 – Specimen Quality of Health Economic Studies (QHES) checklist**

|  | Questions | Points available | Yes | No |
| --- | --- | --- | --- | --- |
| 1. | Was the study objective presented in a clear, specific, and measurable manner? | 7 |  |  |
| 2. | Were the perspective of the analysis (societal, third-party payer, etc.) and reasons for its selection stated? | 4 |  |  |
| 3. | Were variable estimates used in the analysis from the best available source (RCT = best, expert opinion = worst)? | 8 |  |  |
| 4. | If estimates came from a subgroup analysis, were the groups pre-specified at the beginning of the study? | 1 |  |  |
| 5. | Was uncertainty handled by (1) statistical analysis to address random events, (2) sensitivity analysis to cover a range of assumptions? | 9 |  |  |
| 6. | Was incremental analysis performed between alternatives for resources and costs? | 6 |  |  |
| 7. | Was the methodology for data extraction (including the value of health states and other benefits) stated? | 5 |  |  |
| 8. | Did the analytic horizon allow time for all relevant and important outcomes? Were benefits and costs that went beyond 1 year discounted (3% - 5%) and justification given for the discount rate? | 7 |  |  |
| 9. | Was the measurement of costs appropriate and the methodology for the estimation of quantities and unit costs clearly described? | 8 |  |  |
| 10. | Was the primary outcomes measure(s) for the economic evaluation clearly stated and were the major short-term, long-term and negative outcomes included? | 6 |  |  |
| 11. | Were the health outcomes measures/scales valid and reliable? If previously tested valid and reliable measures were not available, was justification given for the measures/scales used? | 7 |  |  |
| 12. | Were the economic model (including structure), study methods and analysis, and the components of the numerator and denominator displayed in a clear, transparent manner? | 8 |  |  |
| 13. | Were the choice of economic model, main assumptions, and limitations of the study stated and justified? | 7 |  |  |
| 14. | Did the author(s) explicitly discuss the direction and magnitude of potential biases? | 6 |  |  |
| 15. | Were the conclusions/recommendations of the study justified and based on the study results? | 8 |  |  |
| 16. | Was there a statement disclosing the source of funding for the study? | 3 |  |  |
| Total score | | 100 |  |  |

**Appendix 4 – Detailed outcomes for all papers**

|  | Hao et al, 2021 | Karlsson et al, 2021 | Hendrix et al, 2021 | Thomas et al, 2021 | Wong et al, 2021 | Callender et al, 2021 | Cenin et al, 2020 | Naber et al, 2019 | Callender, 2019 | Pashayan et al, 2018 |
| --- | --- | --- | --- | --- | --- | --- | --- | --- | --- | --- |
| Study objective | To assess the cost-effectiveness of quadrennial magnetic resonance imaging (MRI)-based screening using either Stockholm3 (S3M) or prostate-specific antigen (PSA) test as a reflex test. | To assess the cost-effectiveness of the Stockholm3 Model (S3M) in screening. | To assess the cost-effectiveness of moving from universal screening (risk- agnostic) to risk-stratified screening | To use risk scores as the basis for determining age at which faecal immunochemical test (FIT) screening should start, then to estimate  the cost-effectiveness, clinical benefits and resource impact of polygenic risk informed -stratification, compared with current screening strategies. | To evaluate the cost-effectiveness of a breast cancer screening programme that incorporates genetic testing against the current biennial mammogram-only screening programme. | To evaluate the benefit, harm and cost-effectiveness of MRI before biopsy compared with biopsy-first screening for prostate cancer using age-based and polygenic risk-stratified screening strategies. | To investigate the impact  of personalizing colorectal cancer screening, based on polygenic risk  and family history and to compare its cost-effectiveness to uniform  screening (using fecal immunochemical testing (FIT) and colonscopies at different intervals and different starting ages) | To assess whether and under what conditions polygenic risk-informed screening for colorectal cancer may be a cost-effective alternative to uniform screening (which involved colonoscopy screening at ages 50,60 and 70 years). | To assess the benefit-harm ratio and cost-effectiveness of a polygenic risk-tailored screening programme for prostate  cancer | To assess the benefit/harm ratio and cost-effectiveness of polygenic risk-stratified breast screening compared with standard age based screening and no screening. |
| Cancer(s) studied | Prostate | Prostate | Prostate | Colorectal | Breast | Prostate | Colorectal | Colorectal | Prostate | Breast |
| Context (screening strategies compared and country) | Swedish setting.  Strategies compared:  No screening  MRI for PSA≥3ng/mL and TBx/SBx for PI-RADS 3-5  MRI for S3M≥15% using a reflex threshold of PSA≥1.5ng/mL and TBx/SBx for men who had PI-RADS 3-5  MRI for S3M≥15% using a reflex threshold of PSA≥2ng/mL and TBx/SBx for men who had PI-RADS 3-5 | Sweden setting.  Strategies compared:  No prostate cancer screening; screening using the PSA test; and screening using the S3M test as a reflex test for PSA values ≥ 1, 1.5 and 2 ng/mL | US setting  Strategies compared:  No screening and nine combinations of starting age (45, 50 or 55) and screening interval (1, 2 or 4 years). Strategies compared for each risk stratum separately. Then compared universal policies to risk-stratified policies in which intermediate-risk  men are screened with the same intensity as in the universal policy and low- and high-risk men receive lower and  higher intensity screening, respectively. | England setting. Biennial faecal immunochemical test (FIT), starting at an age determined through polygenic-informed risk-assessment at age 40, compared to FIT screening that started at a fixed age for all individuals.  Baseline strategy based on current FIT screening strategy in England involving biennial FIT at a threshold of 120µg/g between ages of 60 and 74. Three other comparators involved a start age of 50 instead of 60, the use of threshold of 20µg/g. | Singapore setting. Strategies compared: biennial mammogram with polygenic-risk informed screening. | England setting.  Strategies compared:  No screening  Age-based screening with biopsy if PSA ≥ 3, age-based screening with MRI if PSA ≥ 3 and biopsy if abnormal findings  Risk-stratified screening with PRS as well as age, biopsy if PSA ≥ 3  Risk-stratified screening with PRS, MRI if PSA ≥ 3 and biopsy if abnormal findings | Australia setting  Strategies compared: No screening, plus  25 different  screening strategies defined by different start ages for screening (40, 46, 50, 54, or 60  years), test used (FIT or colonoscopy), and interval (annual, biennial or  triennial screening for FIT, and every 5 or 10 years for colonoscopy). | US setting  Strategies compared:  No screening,  risk-stratified screening based on PRS,  Uniform screening with colonoscopies at ages 50, 60, and 70 years. | England setting  Strategies compared:  no screening,  age-based screening with prostate-specific antigen (PSA) testing  PRS-informed risk-tailored screening | UK setting.  Strategies compared:  No screening,  Age-based screening,  PRS-informed risk stratified screening |
| Type of economic evaluation used | Cost-utility | Cost-utility | Cost-utility | Cost-utility | Cost-utility | Cost-utility | Cost-utility | Cost-utility | Cost-utility | Cost-utility |
| Proposed design for a polygenic risk-informed screening programme | Screening assumed to be administered by general practitioners but otherwise not described. | Not described | Not described | Not described | Assumed that buccal swabs would be collected and used in genotyping. Also asked to complete a questionnaire on breast cancer risk factors, before being stratified into three risk groups based on their initial PRS. Individuals in each risk group will receive their initial PRSs within three to 6 months of a buccal swab. | Not described. | Not described | Assumes that population (at average risk for colorectal cancer) that is willing to undergo polygenic test as well as subsequent risk-informed colonoscopy screening. | Not described. | Not described. |
| Risk thresholds (if used) | Screening based on PSA≥3ng/mL; and S3M risk with reflex test  thresholds of PSA≥1.5 and 2ng/mL. S3M risk prediction based on a genetic risk score based on a single nucleotide polymorphism (SNP) chip,  five plasma protein biomarkers, together with self-reported age, family history and any previous | Men with an S3M risk of Gleason 7 cancer above 10% referred to a urologist.  negative biopsies | Pre-specified thresholds of the Prompt-PGS ©  (Prompt-Prostate Genetic Score (≤0.60, >0.6–1.3, and >1.3) ) used to designate participants as belonging to low-,  intermediate-, and high-risk strata. | Different risk scores were used based on combinations of phenotypic and genetic information. The age for a first FIT invite was calculated as the age at which an individual would reach a specific risk threshold (separately for each risk score) at age 40. This ensured that number of FIT screenings was similar between comparator and intervention screening strategies. | Three groups were defined for the intervention defined by percentile cutoffs of polygenic risk: PRS stratified as <60% as low risk, 60-95th percentile as high risk, and >95% for high risk. Different cutoffs for the risk groups evaluated in a scenario analysis. | Varied the  10-year absolute risk of developing prostate cancer (based on age and polygenic risk) from 2% to 10% in men aged between 55 and 69. | Using previous research, the population was stratified into five risk groups based on quintiles of polygenic risk score. | Each population was split into relative risk groups, into which individuals were assigned based on baseline risk and discriminative accuracy of the polygenic test. | Varied the 10-year absolute risk of developing prostate cancer based on PRS thresholds at which individuals were eligible for screening from 2% to 10% | Risk threshold used in polygenic risk-informed risk based screening in which only the proportion of women in the population with a risk score greater than a threshold risk were offered screening.  Women were screened every three years until age 69 if they met a polygenic-risk informed threshold. 99 scenarios of risk-stratified screening strategies corresponding to percentiles of the risk score were evaluated. |
| Adherence to screening | Assumed entire cohort would participate in screening. | Assumed entire cohort would participate in screening. | Assumed entire cohort would participate in screening. | One screening scenario reduced modelled screening update by 25% to account for uncertainty of the impact of risk stratification, and otherwise screening uptake is assumed to be unaffected by risk stratification although update varies by age, sex, deprivation and prior response to screening. | Assumed entire cohort would participate in screening. | Assumed entire cohort would participate in screening in base case analysis. Sensitivity analysis varied uptake of both PSA and polygenic risk stratification to 75%. | Base case analysis assumed perfect adherence.  Subsequently estimated costs and effects of screening at adherence levels currently observed in Australia. | In base-case analysis, assumed full adherence to polygenic testing, colonoscopy screening, and colonoscopy surveillance. Sensitivity analyses modelled observed adherence rates in the US | Assumed entire cohort would participate in screening in base case analysis. Sensitivity analysis varied uptake of both PSA and polygenic risk stratification to 75%. | Assumed entire cohort would participate in screening in base case analysis. In sensitivity analysis examined impact of 90% and 75% adherence to the screening  recommendation for higher and lower risk groups. |
| Screening interval modelled | Quadrennial between ages of 55 and 69. | Quadrennial between ages of 55 and 69 | Annual, biennial and quadrennial. | Biennial, reflecting current FIT screening strategy in England. | Biennial screening for conventional mammogram screening. Polygenic-risk informed screening may comprise self-examination, or annual, biennial or triennial screening depending on risk score. | Quadrennial between ages of 55 and 69. | Dependent on technology. Annual, biennial or triennial screening for FIT, and quinquennial or decennial for colonoscopy | Screening intervals from 1 to 20 years modelled (at intervals of 1, 2, 3, 5, 7, 10, 15, and 20 years.) | Quadrennial age-based PS from 55 to 69 under age-based screening, and quadrennial PSA testing when risk threshold reached for men aged 55 to 69 when under PRS-informed risk-tailored screening | Triennial from age 50 to 69 under both age-based screening and PRS-informed risk-based screening once risk threshold was met. |
| Structure of the model | Microsimulation model (Prostata model) | Microsimulation model (Prostata model) | Microsimulation model (Fred Hutchinson Cancer Research Centre model) | Microsimulation model (MiMiC-Bowel). | Markov model | Life table cohort Markov model | Microsimulation model (the MISCAN-Colon model) | Microsimulation model (the MISCAN-Colon model) | Life table cohort Markov model | Lifetable cohort model |
| Age range of cohort | From age 55 and followed to remainder of lifetime. | From birth and followed over lifetime. | 40 years of age (with different screening start ages >40) and followed until age 100. Screening assumed to stop at age 69. | 30 and over. Screening takes place at various ages depending on strategy. Risk-assessment assumed to be carried out at age 40. | Women aged between 35 and 74 | Screening took place at 55-69 years of age with follow-up to 90 years of age | 40 years of age (and born in 1980) and followed until age 100, at which point individuals in the cohort were assumed to be dead. Screening assumed to stop at age 74. | 40 years of age with US life expectancy, and followed until death. Screening modelled as ending between 70 and 85 years of age. | Screening took place at 55-69 years of age with follow-up to 90 years of age | 50 years of age with follow-up to 85 years. |
| Size of cohort modelled | 10 million. | Not directly stated but references related work which refers to cohorts of 100m men | 100 million | 6,787,000 | 3, 014,388 individuals included in models. Not otherwise reported | 4.48 million | 100 million | Polygenic risk cohort described as consisting of >1m simulated individuals. Not otherwise reported. | 4.48 million | 364,500 |
| Perspective of the analysis | Both societal and healthcare perspectives | Societal perspective | Not explicitly stated but only health system costs included in analysis | Health system perspective | Health system perspective | Health system perspective | Health system perspective | Modified societal perspective comprising direct medical costs and time costs for patients and patient escorts. Non-health care costs such as travel costs were not included. | Health system perspective | Health system perspective |
| Cancer treatments modelled | Active surveillance, radical prostatectomy, radiation therapy | Active surveillance, radical prostatectomy and radiation therapy | Primary surgery, radiation therapy, active surveillance for low grade cancer, second-generation androgen receptor  inhibitors for distant stage cancer | Treatments defined by stage of cancer. Patients found to have adenomas undergo polypectomy and applicable guidelines are implemented in the model for surveillance following adenoma removal | Treatments not specified. | Active surveillance, radical prostatectomy, radical radiotherapy, brachytherapy, chemotherapy, androgen deprivation therapy. | Treatments defined by stage and location of cancer in Australian cancer care. | Treatments defined by stage of cancer, but not otherwise specified. | Treatments based on the National Institute for Health and Care Excellence (NICE) prostate cancer pathway and NICE prostate cancer guideline. Active surveillance, radical prostatectomy, radical radiotherapy, brachytherapy, chemotherapy, androgen deprivation therapy. | Treatment of primary breast cancer and treatment of advanced metastatic cancer, but not otherwise specified. |
| Modelling of cancer progression | Progression modelled between preclinical states (T1-T2, T3-T4 and metastasis) | Pre-clinical progression for T- and M-stage for a given Gleason score prior to diagnosis modelled. | Progression from localized to metastatic prostate cancer within Gleason grades 2-6, 7, or 8-10 modelled as well as progression from preclinical to clinical states. | Patients assumed to have normal colorectal epithelium at age 30 and then transition through nine possible states ranging from healthy epithelium to low and high risk adenoma, colorectal stages A to D , death from colorectal cancer or death from other causes. Serrated adenoma pathway modelled by transition directly from normal epithelium to CRC stage A. | Patients can transition from the healthy state to breast cancer stages I-IV or death. Patients cannot experience remission and do not transition between cancer stages. | Cancer progression not explicitly modelled | Natural history modelling of cancer progression (no lesion to screen-detectable adenoma phase (based on size of adenoma), screen detectable cancer phase (stages I to IV), to clinical colorectal stages (stages I to IV). As each simulated  person ages, one or more adenomas may arise and some can progress  in size from small (<5 mm) to medium (6–9 mm) to large (>10 mm). | Adenomas can progress  from small (5 mm), to medium (6–9 mm), to large size (10 mm). Some adenomas can develop into cancer,  which may progress through preclinical and clinical colorectal cancer stages I to IV. | Cancer progression not modelled due to uncertainty about transition rates between states and about impact of polygenic risk on these transitions | Not explicitly modelled, although model accounts for primary breast cancer and advanced metastatic breast cancer. |
| Mortality measures considered | Prostate cancer death and death from other causes | Prostate cancer death and death from other causes | Prostate cancer death and death from other causes. | Colorectal cancer deaths and deaths from other causes | Breast cancer death and deaths from other causes. | Prostate cancer death and death from other causes | Colorectal cancer death and death from all other causes | Colorectal cancer death and death from all other causes | Prostate cancer death and death from all other causes | Breast cancer death and death from all other causes |
| Health state utility values considered (for example the sources used and any anxiety associated with a high-risk PRS diagnosis) | Based on general population health values Utility decrement assigned to PSA test, biopsy, cancer diagnosis, treatment/active surveillance, metastatic disease, post-recovery, palliative therapy and terminal illness. Potential psychological impacts of screening are not included | Based on general population health values Utility decrement assigned to PSA test, biopsy, cancer diagnosis, treatment/active surveillance, post-recovery, palliative therapy and terminal illness. Potential psychological impacts of screening are not included | The “healthy state” was assigned a value of 1.0. There was no adjustment for age or for any utility impact from screening. Decrements were applied for biopsy, surveillance, treatment, symptomatic cancer and distant and end-of life states. | Age and sex-adjusted population figures were adjusted by adjustments for colorectal cancer stages. Adjustments were made for bowel perforation and intestinal bleed. Assumed no disutility from determining or knowing polygenic risk score. | Stage-specific utility values were calculated from Wong et al. The “healthy state” was assigned a value of 1.0. There was no adjustment for age or for any utility impact from determining or knowing polygenic risk score. | Age-adjusted utility values from the general population. Specific reduction in utility only for those with prostate cancer. | Source or level of background utility values not described. Assumed utility loss due to  (colorectal cancer) screening colonoscopy,  complication of colonoscopy and colorectal cancer care. Assumed no disutility from determining or knowing polygenic risk score. | Source or level of background utility values not described. Assumed utility loss due to  colonoscopy,  complications of colonoscopy and colorectal cancer care by stage. Assumed no disutility of obtaining or knowing polygenic risk. | Background age-specific utility estimates, modified where necessary by a prostate treatment value and for post-treatment recovery. Assumed no disutility of obtaining or knowing polygenic risk. | Background age-specific utility estimates, modified where necessary by a utility decrement associated with a diagnosis of breast cancer. Assumed no disutility of obtaining or knowing polygenic risk. |
| Duration of follow-up modelled | Lifetime, from age 55. | Lifetime, from birth. | 60 years from 40 to 100 years of age. | Lifetime follow-up. | 40 years of follow-up until age 74 | 35 years of follow-up, or until age 90 years, whichever was first | 60 years of follow-up until death or 100 years of age | Lifetime follow-up, starting at 40 years of age. | 35 years of follow-up, or until age 90 years, whichever was first | 35 years of follow up from 50 years to 85 years of age. |
| Outcome measure (for example cost per Quality Adjusted Life Year gained) | Costs per quality-adjusted life year. | Costs per quality-adjusted life year. | Costs per quality-adjusted life year. | Costs per quality adjusted life year. | Costs per quality adjusted life year. | Costs per quality-adjusted life year | Costs per quality-adjusted life year | Costs per quality-adjusted life year | Costs per quality-adjusted life year | Costs per quality-adjusted life year |
| How are genetic data obtained (or assumed to be obtained) and analyzed? | Not reported – assumed genetic data available for all men in cohort | Not reported – assumed genetic data available for all men in cohort | Not reported – assumed genetic data available for all individuals in cohort | Not reported. Risk assessment assumed to be carried out in all modelled individuals at age 40. Method of assessment not reported. | Individuals genotyped by buccal swab but not otherwise specified. | Not reported – assumed genetic data available for all men in cohort | Assumed that risk was determined by an assessment for family history and polygenic testing prior to screening. Assumed colorectal cancer family history would be taken by a general practitioner. | Not reported – assumed genetic data available for all individuals in cohort | Not reported – assumed genetic data available for all men in cohort | Not reported - assumed genetic data available for all women in cohort |
| Assumptions made in creating the polygenic risk score | Used the  Stockholm3 (S3M) risk-model that combines PSA, SNPs and other established and new plasma biomarkers.  The polygenic risk score is based on 232 SNPs (Gronberg et al) | Used the  Stockholm3 (S3M) risk-model that combines PSA, SNPs and other established and new plasma biomarkers. The polygenic risk score is based on 232 SNPs (Gronberg et al) | The Prompt-PGS® risk score is a weighted count of Prostate cancer risk-associated single nucleotide polymorphism alleles, where the weights reflect both the odds ratio of Prostate cancer diagnosis and the allele frequency in a population. | Based on 120 risk colorectal cancer risk alleles identified in Huyghe et al | A polygenic risk score was not used. Instead, percentiles of a an otherwise unspecified polygenic risk distribution were used to create low, medium and high risk groups. Polygenic risk was modelled as a “multiplier” that influenced higher or lower transitions from the healthy state to cancer depending on risk group membership. | Based on 175 prostate cancer susceptibility loci identified in genome-wide association studies. Loci assumed to interact log additively to define a lognormal distribution of polygenic risk in the population on a relative risk scale. | Based on 45 SNPs known to increase risk of colorectal cancer | Created a hypothetical population with individual-specific risk to which genetic variants were added until the area under the curve (AUC) of a polygenic test reached a pre-specified value. This predicted risk was then divided by population prevalence to create a relative risk, which was categorized into 60 groups. | Number of loci not specified but based on Schumacher et al., and Dadaev, et al. Loci assumed to interact log additively to define a lognormal distribution of polygenic risk in the population on a relative risk scale. | Based on 310 known breast cancer susceptibility loci. |
| Cost of the PRS and any associated costs | €251 including PSA test analysis, GP visit and S3M test analysis | €255 (S3M test including GP visit) | $250. Based on commercial costs of the Prompt-PGS software. | No costs assigned to risk scoring. Instead, cost analysis carried out to determine maximum justifiable cost of implementing risk-scoring in population at age 40. | Genotyping of buccal swab assumed to cost SGD210. | £25. Based on personal communication of tariffs used in the English National Health Service. | Assumed cost ($200) based on a commercially available polygenic test for breast cancer | Assumed cost ($200) based on currently available commercial polygenic tests. | £25. Estimated from personal discussion of costs charged to NHS hospitals for prostate cancer genome wide associations studies. | £50. Based on per variant  research cost of genotyping |
| How were PRS data included and modelled? | A positive S3M test was defined as one having a PSA value above the reflex threshold and a risk prediction above 10%. | A positive S3M test was defined as one having a PSA value above the reflex threshold and a risk prediction above 10%. | Estimated hazard ratios for incidence within strata of risk defined by pre-specified Prompt-PGS risk scores to identify those at low, intermediate and high risk relative to the average risk population. | Each modelled individual was randomly assigned risk alleles to reflect allele frequency in UK Biobank data, and accounting for correlations between alleles on same chromosome. | Individuals were stratified into three risk groups  based on their initial PRS – low, intermediate, and high. The PRSs are stratified by setting cutoffs at below 60^th^ percentile for the low-risk group, 60th to 95th percentile  for the intermediate-risk group and above 95th percentile for the high-risk group.  Transition probabilities between health and disease states influenced by “multipliers” of 2x, 1x ,and 0.5x for each group. | From log relative risk distribution derived the age-specific proportion of men above each 10-year absolute risk threshold, and proportion of all cancers that would be diagnosed in these men. These proportions were used to calculate the  age-specific relative risk of developing prostate cancer in those men above and below the 10-year absolute risk thresholds. | Relative risk (compared to population average risk) of developing colorectal cancer was based on a combination of family history and quintile of PRS distribution. Family history and quintile of PRS risk were observed to be largely independent. | A relative risk distribution was generated in hypothetical populations with varying area under the curve (AUC) values of polygenic testing of  0.60, 0.65, 0.70, 0.75, and 0.80. This population was split into groups of estimated relative risk, which assigned individuals to a relative risk group based on baseline risk and accuracy of the polygenic test. | From log relative risk of prostate cancer for each risk threshold relative to the background 10-year absolute risk of developing this cancer in the absence of screening. The log relative risk of developing prostate cancer was then applied to the polygenic risk distribution to determine the proportion of cases above the threshold. This was used to derive the relative risk of developing prostate cancer amongst the screened and unscreened. | Assumed log-additive interactions between genetic and other risk factors to obtain a log-normal distribution of risk on the relative risk scale.  Percentile rank of relative risk or (age-conditional) absolute risk was calculated.  Calculated the  relative risk associated with a risk score in higher- and lower-risk subgroups in relation to a relative risk distribution. |
| Whether ethnicity was considered in relation to PRS, and whether differential cost-effectiveness was considered by ethnicity | Ethnicity not considered | Ethnicity not considered | Ethnicity not considered. | Ethnicity was included as a phenotypic risk factor. Differential cost-effectiveness by ethnicity not considered. | Percentile risk group definitions were adjusted to account for Asian ancestry. Ethnicity not otherwise considered. | Ethnicity not considered. | Ethnicity not considered. | Not explicitly modelled, although some discussion of how adherence may vary by ethnicity | Ethnicity not considered. | Ethnicity not considered. |
| Cost-effectiveness threshold used | €47,218  (SEK 500,000) per QALY | €0 per QALY and €50,000 per QALY | A formal ex ante cost-effectiveness threshold not used – instead, strategies compared on the basis of incremental cost-effectiveness ratios | £20,000 and £30,000 per QALY both used as cost-effectiveness thresholds. | An ex-ante cost-effectiveness threshold was not used. Different thresholds were calculated to assess the probability of polygenic risk-informed screening being cost-effective. | £20,000 and £30,000 per QALY | AUS$50,000 per QALY | Cost-effectiveness threshold for PRS-informed risk stratification was set equal to level at which QALYs gained were equivalent to those of uniform screening. $69,000, $65,000, $56,700, $46,000, and $38,500 for AUC of 0.60, 0.65, 0.70, 0.75, and 0.80, respectively | £20,000 and £30,000 per QALY | £20,000 and £30,000 per QALY |
| Cost-effectiveness results of PRS-informed screening compared to non-PRS screening modalities | Stockholm3 with a reflex threshold of PSA≥2ng/mL had the lowest ICER,  €38,894 per QALY gained, in the base case analysis | Prostate cancer screening using the polygenic risk-informed S3M test for men with an initial PSA ≥ 2.0 ng/mL was cost-effective compared with screening using only PSA. | Cost-effectiveness of PRS-informed risk screening compared to universal screening depended on universal screening policy modelled. PRS informed risk-stratified screening most likely to be cost-effective when universal screening is performed on an annual basis starting at age 55. | PRS-informed screening was very likely to be cost-effective when used in conjunction with phenotypic information compared to screening strategies relying on phenotypic data alone. | Compared with biennial mammogram-only screening, polygenic-risk informed screening had lower costs and higher quality-adjusted life years and was very likely to be cost-effective. | MRI-first risk-stratified screening scenarios at risk thresholds ≥3.5% were more cost-effective than no screening at a cost-effectiveness threshold of £20,000. Strategies with highest net monetary benefit at cost-effectiveness thresholds of £20,000 and £30,000 were MRI-first risk-stratified screening at risk thresholds of 8.5% and 7.5%, respectively | Uniform screening was more likely to be cost-effective than PRS-informed risk-based screening. Personalized and uniform screening scenarios yielded similar QALYs. Personalized screening cost more than uniform screening, largely due to the cost of determining risk. | Polygenic risk-informed unlikely to be cost-effective; this form of screening yielded same number of QALYs as uniform screening at increased costs. | Risk-based screening was cost-effective at a cost-effectiveness threshold of £20,000 per QALY gained  compared to no screening at all 10-year absolute risk thresholds above 4.5%. At all 10-year absolute risk < 10%, risk-based screening led to a greater number of incremental QALYs gained than age-based screening  whilst incurring fewer additional costs at all risk thresholds above 2%. | PRS-informed risk stratification at the 70^th^ percentile had the highest net monetary benefit, with a 72% probability of being cost-effective at a at a cost-effectiveness threshold of £20,000. |
| Sensitivity of cost-effectiveness results to model parameters. | Reducing the unit cost of Stockholm3 to €94 (57% reduction)  resulted in a 16% reduction in the ICER.  Results sensitive to the discount rates used. | The S3M test was more cost effective at higher  reflex thresholds, at higher biopsy costs and at lower S3M test costs | The cost of the polygenic risk scoring, biopsy and surveillance influence the relative cost-effectiveness compared to universal age-based screening. | Risk assessment costs >£114 would not be cost effective. Cost-effectiveness was lower when risk scores were less discriminatory, with lower mean start ages for screening, or higher FIT thresholds. Men were more likely to benefit from risk-stratified screening than women. | Results were not sensitive to several key model parameters including the low- and high-risk multipliers, direct medical costs for Stage II breast cancer,  and the sensitivity of mammogram and ultrasound tests. | MRI-first risk-stratified screening strategies were sensitive to the cost of polygenic risk stratification (varied from £25 to £100). MRI-first risk-stratified screening was insensitive to a 75% (baseline 100%) uptake of  polygenic risk stratification. | Results were sensitive to the cost of determining polygenic risk. A threshold analysis found that the costs of determining polygenic risk should not exceed $47.52 for risk-informed screening to be cost-effective at a cost-effectiveness threshold of $50,000. | Risk-stratified screening could be considered cost-effective if polygenic testing costs were 30% less expensive, if the AUC of polygenic testing increased by 0.05, or a greater than 5% increase in screening adherence. | Risk-stratified screening strategies were somewhat sensitive to the cost of polygenic risk stratification (varied from £25 to £50) and to incomplete adherence. | Polygenic-informed risk stratification was somewhat less likely to be cost effective the higher the cost of the risk assessment and the lower the levels of adherence. |

**Appendix 5 – Completed QHES checklist for all included studies**

**Quality of Health Economic Studies (QHES) instrument – Hao et al 2021**

|  | Questions | Points available | Yes | No |
| --- | --- | --- | --- | --- |
| 1. | Was the study objective presented in a clear, specific, and measurable manner? | 7 | x |  |
| 2. | Were the perspective of the analysis (societal, third-party payer, etc.) and reasons for its selection stated? | 4 | x |  |
| 3. | Were variable estimates used in the analysis from the best available source (RCT = best, expert opinion = worst)? | 8 | x |  |
| 4. | If estimates came from a subgroup analysis, were the groups pre-specified at the beginning of the study? | 1 | x (not applicable) |  |
| 5. | Was uncertainty handled by (1) statistical analysis to address random events, (2) sensitivity analysis to cover a range of assumptions? | 9 | x |  |
| 6. | Was incremental analysis performed between alternatives for resources and costs? | 6 | x |  |
| 7. | Was the methodology for data extraction (including the value of health states and other benefits) stated? | 5 | x |  |
| 8. | Did the analytic horizon allow time for all relevant and important outcomes? Were benefits and costs that went beyond 1 year discounted (3% - 5%) and justification given for the discount rate? | 7 | x |  |
| 9. | Was the measurement of costs appropriate and the methodology for the estimation of quantities and unit costs clearly described? | 8 | x |  |
| 10. | Was the primary outcomes measure(s) for the economic evaluation clearly stated and were the major short-term, long-term and negative outcomes included? | 6 |  | x |
| 11. | Were the health outcomes measures/scales valid and reliable? If previously tested valid and reliable measures were not available, was justification given for the measures/scales used? | 7 | x |  |
| 12. | Were the economic model (including structure), study methods and analysis, and the components of the numerator and denominator displayed in a clear, transparent manner? | 8 | x |  |
| 13. | Were the choice of economic model, main assumptions, and limitations of the study stated and justified? | 7 | x |  |
| 14. | Did the author(s) explicitly discuss the direction and magnitude of potential biases? | 6 | x |  |
| 15. | Were the conclusions/recommendations of the study justified and based on the study results? | 8 | x |  |
| 16. | Was there a statement disclosing the source of funding for the study? | 3 | x |  |
| Total score | | 100 | 94 |  |

**Quality of Health Economic Studies (QHES) instrument – Karlsson et al 2021**

|  | Questions | Points available | Yes | No |
| --- | --- | --- | --- | --- |
| 1. | Was the study objective presented in a clear, specific, and measurable manner? | 7 | x |  |
| 2. | Were the perspective of the analysis (societal, third-party payer, etc.) and reasons for its selection stated? | 4 | x |  |
| 3. | Were variable estimates used in the analysis from the best available source (RCT = best, expert opinion = worst)? | 8 | x |  |
| 4. | If estimates came from a subgroup analysis, were the groups pre-specified at the beginning of the study? | 1 | x (not applicable) |  |
| 5. | Was uncertainty handled by (1) statistical analysis to address random events, (2) sensitivity analysis to cover a range of assumptions? | 9 | x |  |
| 6. | Was incremental analysis performed between alternatives for resources and costs? | 6 | x |  |
| 7. | Was the methodology for data extraction (including the value of health states and other benefits) stated? | 5 | x |  |
| 8. | Did the analytic horizon allow time for all relevant and important outcomes? Were benefits and costs that went beyond 1 year discounted (3% - 5%) and justification given for the discount rate? | 7 | x |  |
| 9. | Was the measurement of costs appropriate and the methodology for the estimation of quantities and unit costs clearly described? | 8 | x |  |
| 10. | Was the primary outcomes measure(s) for the economic evaluation clearly stated and were the major short-term, long-term and negative outcomes included? | 6 |  | x |
| 11. | Were the health outcomes measures/scales valid and reliable? If previously tested valid and reliable measures were not available, was justification given for the measures/scales used? | 7 | x |  |
| 12. | Were the economic model (including structure), study methods and analysis, and the components of the numerator and denominator displayed in a clear, transparent manner? | 8 | x |  |
| 13. | Were the choice of economic model, main assumptions, and limitations of the study stated and justified? | 7 | x |  |
| 14. | Did the author(s) explicitly discuss the direction and magnitude of potential biases? | 6 | x |  |
| 15. | Were the conclusions/recommendations of the study justified and based on the study results? | 8 | x |  |
| 16. | Was there a statement disclosing the source of funding for the study? | 3 | x |  |
| Total score | | 100 | 94 |  |

**Quality of Health Economic Studies (QHES) instrument – Hendrix et al 2021**

|  | Questions | Points available | Yes | No |
| --- | --- | --- | --- | --- |
| 1. | Was the study objective presented in a clear, specific, and measurable manner? | 7 | x |  |
| 2. | Were the perspective of the analysis (societal, third-party payer, etc.) and reasons for its selection stated? | 4 | x |  |
| 3. | Were variable estimates used in the analysis from the best available source (RCT = best, expert opinion = worst)? | 8 | x |  |
| 4. | If estimates came from a subgroup analysis, were the groups pre-specified at the beginning of the study? | 1 | x (not applicable) |  |
| 5. | Was uncertainty handled by (1) statistical analysis to address random events, (2) sensitivity analysis to cover a range of assumptions? | 9 | x |  |
| 6. | Was incremental analysis performed between alternatives for resources and costs? | 6 | x |  |
| 7. | Was the methodology for data extraction (including the value of health states and other benefits) stated? | 5 |  | x (choice of perfect health for the healthy state not justified in paper) |
| 8. | Did the analytic horizon allow time for all relevant and important outcomes? Were benefits and costs that went beyond 1 year discounted (3% - 5%) and justification given for the discount rate? | 7 | x |  |
| 9. | Was the measurement of costs appropriate and the methodology for the estimation of quantities and unit costs clearly described? | 8 | x |  |
| 10. | Was the primary outcomes measure(s) for the economic evaluation clearly stated and were the major short-term, long-term and negative outcomes included? | 6 |  | x |
| 11. | Were the health outcomes measures/scales valid and reliable? If previously tested valid and reliable measures were not available, was justification given for the measures/scales used? | 7 | x |  |
| 12. | Were the economic model (including structure), study methods and analysis, and the components of the numerator and denominator displayed in a clear, transparent manner? | 8 | x |  |
| 13. | Were the choice of economic model, main assumptions, and limitations of the study stated and justified? | 7 | x |  |
| 14. | Did the author(s) explicitly discuss the direction and magnitude of potential biases? | 6 | x |  |
| 15. | Were the conclusions/recommendations of the study justified and based on the study results? | 8 | x |  |
| 16. | Was there a statement disclosing the source of funding for the study? | 3 | x |  |
| Total score | | 100 | 89 |  |

**Quality of Health Economic Studies (QHES) instrument – Thomas et al 2021**

|  | Questions | Points available | Yes | No |
| --- | --- | --- | --- | --- |
| 1. | Was the study objective presented in a clear, specific, and measurable manner? | 7 | x |  |
| 2. | Were the perspective of the analysis (societal, third-party payer, etc.) and reasons for its selection stated? | 4 | x |  |
| 3. | Were variable estimates used in the analysis from the best available source (RCT = best, expert opinion = worst)? | 8 | x |  |
| 4. | If estimates came from a subgroup analysis, were the groups pre-specified at the beginning of the study? | 1 | x (not applicable) |  |
| 5. | Was uncertainty handled by (1) statistical analysis to address random events, (2) sensitivity analysis to cover a range of assumptions? | 9 | x |  |
| 6. | Was incremental analysis performed between alternatives for resources and costs? | 6 | x |  |
| 7. | Was the methodology for data extraction (including the value of health states and other benefits) stated? | 5 | x |  |
| 8. | Did the analytic horizon allow time for all relevant and important outcomes? Were benefits and costs that went beyond 1 year discounted (3% - 5%) and justification given for the discount rate? | 7 | x |  |
| 9. | Was the measurement of costs appropriate and the methodology for the estimation of quantities and unit costs clearly described? | 8 | x |  |
| 10. | Was the primary outcomes measure(s) for the economic evaluation clearly stated and were the major short-term, long-term and negative outcomes included? | 6 |  | x |
| 11. | Were the health outcomes measures/scales valid and reliable? If previously tested valid and reliable measures were not available, was justification given for the measures/scales used? | 7 | x |  |
| 12. | Were the economic model (including structure), study methods and analysis, and the components of the numerator and denominator displayed in a clear, transparent manner? | 8 | x |  |
| 13. | Were the choice of economic model, main assumptions, and limitations of the study stated and justified? | 7 | x |  |
| 14. | Did the author(s) explicitly discuss the direction and magnitude of potential biases? | 6 | x |  |
| 15. | Were the conclusions/recommendations of the study justified and based on the study results? | 8 | x |  |
| 16. | Was there a statement disclosing the source of funding for the study? | 3 | x |  |
| Total score | | 100 | 94 |  |

**Quality of Health Economic Studies (QHES) instrument – Wong et al 2021**

|  | Questions | Points available | Yes | No |
| --- | --- | --- | --- | --- |
| 1. | Was the study objective presented in a clear, specific, and measurable manner? | 7 | x |  |
| 2. | Were the perspective of the analysis (societal, third-party payer, etc.) and reasons for its selection stated? | 4 | x |  |
| 3. | Were variable estimates used in the analysis from the best available source (RCT = best, expert opinion = worst)? | 8 | x |  |
| 4. | If estimates came from a subgroup analysis, were the groups pre-specified at the beginning of the study? | 1 | x (not applicable) |  |
| 5. | Was uncertainty handled by (1) statistical analysis to address random events, (2) sensitivity analysis to cover a range of assumptions? | 9 | x |  |
| 6. | Was incremental analysis performed between alternatives for resources and costs? | 6 | x |  |
| 7. | Was the methodology for data extraction (including the value of health states and other benefits) stated? | 5 |  | X (choice of perfect health for the healthy state not justified in paper) |
| 8. | Did the analytic horizon allow time for all relevant and important outcomes? Were benefits and costs that went beyond 1 year discounted (3% - 5%) and justification given for the discount rate? | 7 | x |  |
| 9. | Was the measurement of costs appropriate and the methodology for the estimation of quantities and unit costs clearly described? | 8 |  | x (costs are from an unpublished PhD thesis by lead author and are not otherwise described) |
| 10. | Was the primary outcomes measure(s) for the economic evaluation clearly stated and were the major short-term, long-term and negative outcomes included? | 6 |  | x |
| 11. | Were the health outcomes measures/scales valid and reliable? If previously tested valid and reliable measures were not available, was justification given for the measures/scales used? | 7 | x |  |
| 12. | Were the economic model (including structure), study methods and analysis, and the components of the numerator and denominator displayed in a clear, transparent manner? | 8 |  | X (no remission or transition between states modelled) |
| 13. | Were the choice of economic model, main assumptions, and limitations of the study stated and justified? | 7 |  | X (no justification for not modelling remission or progression between cancer stages) |
| 14. | Did the author(s) explicitly discuss the direction and magnitude of potential biases? | 6 | x |  |
| 15. | Were the conclusions/recommendations of the study justified and based on the study results? | 8 | x |  |
| 16. | Was there a statement disclosing the source of funding for the study? | 3 | x |  |
| Total score | | 100 | 66 |  |

**Quality of Health Economic Studies (QHES) instrument – Callender et al 2021**

|  | Questions | Points available | Yes | No |
| --- | --- | --- | --- | --- |
| 1. | Was the study objective presented in a clear, specific, and measurable manner? | 7 | x |  |
| 2. | Were the perspective of the analysis (societal, third-party payer, etc.) and reasons for its selection stated? | 4 | x |  |
| 3. | Were variable estimates used in the analysis from the best available source (RCT = best, expert opinion = worst)? | 8 | x |  |
| 4. | If estimates came from a subgroup analysis, were the groups pre-specified at the beginning of the study? | 1 | x (not applicable) |  |
| 5. | Was uncertainty handled by (1) statistical analysis to address random events, (2) sensitivity analysis to cover a range of assumptions? | 9 | x |  |
| 6. | Was incremental analysis performed between alternatives for resources and costs? | 6 | x |  |
| 7. | Was the methodology for data extraction (including the value of health states and other benefits) stated? | 5 | x |  |
| 8. | Did the analytic horizon allow time for all relevant and important outcomes? Were benefits and costs that went beyond 1 year discounted (3% - 5%) and justification given for the discount rate? | 7 | x |  |
| 9. | Was the measurement of costs appropriate and the methodology for the estimation of quantities and unit costs clearly described? | 8 | x |  |
| 10. | Was the primary outcomes measure(s) for the economic evaluation clearly stated and were the major short-term, long-term and negative outcomes included? | 6 |  | x |
| 11. | Were the health outcomes measures/scales valid and reliable? If previously tested valid and reliable measures were not available, was justification given for the measures/scales used? | 7 | x |  |
| 12. | Were the economic model (including structure), study methods and analysis, and the components of the numerator and denominator displayed in a clear, transparent manner? | 8 | x |  |
| 13. | Were the choice of economic model, main assumptions, and limitations of the study stated and justified? | 7 | x |  |
| 14. | Did the author(s) explicitly discuss the direction and magnitude of potential biases? | 6 | x |  |
| 15. | Were the conclusions/recommendations of the study justified and based on the study results? | 8 | x |  |
| 16. | Was there a statement disclosing the source of funding for the study? | 3 |  | x (discloses conflicts but not explicitly specific funding) |
| Total score | | 100 | 91 |  |

**Quality of Health Economic Studies (QHES) instrument – Cenin et al, 2020**

|  | Questions | Points available | Yes | No |
| --- | --- | --- | --- | --- |
| 1. | Was the study objective presented in a clear, specific, and measurable manner? | 7 | x |  |
| 2. | Were the perspective of the analysis (societal, third-party payer, etc.) and reasons for its selection stated? | 4 | x |  |
| 3. | Were variable estimates used in the analysis from the best available source (RCT = best, expert opinion = worst)? | 8 | x |  |
| 4. | If estimates came from a subgroup analysis, were the groups pre-specified at the beginning of the study? | 1 | x (not applicable) |  |
| 5. | Was uncertainty handled by (1) statistical analysis to address random events, (2) sensitivity analysis to cover a range of assumptions? | 9 | x |  |
| 6. | Was incremental analysis performed between alternatives for resources and costs? | 6 | x |  |
| 7. | Was the methodology for data extraction (including the value of health states and other benefits) stated? | 5 |  | x (Baseline utility data and sources not reported) |
| 8. | Did the analytic horizon allow time for all relevant and important outcomes? Were benefits and costs that went beyond 1 year discounted (3% - 5%) and justification given for the discount rate? | 7 | x |  |
| 9. | Was the measurement of costs appropriate and the methodology for the estimation of quantities and unit costs clearly described? | 8 | x |  |
| 10. | Was the primary outcomes measure(s) for the economic evaluation clearly stated and were the major short-term, long-term and negative outcomes included? | 6 |  | x |
| 11. | Were the health outcomes measures/scales valid and reliable? If previously tested valid and reliable measures were not available, was justification given for the measures/scales used? | 7 | x |  |
| 12. | Were the economic model (including structure), study methods and analysis, and the components of the numerator and denominator displayed in a clear, transparent manner? | 8 | x |  |
| 13. | Were the choice of economic model, main assumptions, and limitations of the study stated and justified? | 7 | x |  |
| 14. | Did the author(s) explicitly discuss the direction and magnitude of potential biases? | 6 | x |  |
| 15. | Were the conclusions/recommendations of the study justified and based on the study results? | 8 | x |  |
| 16. | Was there a statement disclosing the source of funding for the study? | 3 | x |  |
| Total score | | 100 | 89 |  |

**Quality of Health Economic Studies (QHES) instrument – Naber et al, 2019**

|  | Questions | Points available | Yes | No |
| --- | --- | --- | --- | --- |
| 1. | Was the study objective presented in a clear, specific, and measurable manner? | 7 | x |  |
| 2. | Were the perspective of the analysis (societal, third-party payer, etc.) and reasons for its selection stated? | 4 | x |  |
| 3. | Were variable estimates used in the analysis from the best available source (RCT = best, expert opinion = worst)? | 8 | x |  |
| 4. | If estimates came from a subgroup analysis, were the groups pre-specified at the beginning of the study? | 1 | x |  |
| 5. | Was uncertainty handled by (1) statistical analysis to address random events, (2) sensitivity analysis to cover a range of assumptions? | 9 | x |  |
| 6. | Was incremental analysis performed between alternatives for resources and costs? | 6 | x |  |
| 7. | Was the methodology for data extraction (including the value of health states and other benefits) stated? | 5 |  | x (Baseline utility data and sources not reported) |
| 8. | Did the analytic horizon allow time for all relevant and important outcomes? Were benefits and costs that went beyond 1 year discounted (3% - 5%) and justification given for the discount rate? | 7 | x |  |
| 9. | Was the measurement of costs appropriate and the methodology for the estimation of quantities and unit costs clearly described? | 8 | x |  |
| 10. | Was the primary outcomes measure(s) for the economic evaluation clearly stated and were the major short-term, long-term and negative outcomes included? | 6 |  | x |
| 11. | Were the health outcomes measures/scales valid and reliable? If previously tested valid and reliable measures were not available, was justification given for the measures/scales used? | 7 | x |  |
| 12. | Were the economic model (including structure), study methods and analysis, and the components of the numerator and denominator displayed in a clear, transparent manner? | 8 | x |  |
| 13. | Were the choice of economic model, main assumptions, and limitations of the study stated and justified? | 7 | x |  |
| 14. | Did the author(s) explicitly discuss the direction and magnitude of potential biases? | 6 | x |  |
| 15. | Were the conclusions/recommendations of the study justified and based on the study results? | 8 | x |  |
| 16. | Was there a statement disclosing the source of funding for the study? | 3 | x |  |
| Total score | | 100 | 89 |  |

**Quality of Health Economic Studies (QHES) instrument – Callender et al 2019**

|  | Questions | Points available | Yes | No |
| --- | --- | --- | --- | --- |
| 1. | Was the study objective presented in a clear, specific, and measurable manner? | 7 | x |  |
| 2. | Were the perspective of the analysis (societal, third-party payer, etc.) and reasons for its selection stated? | 4 | x |  |
| 3. | Were variable estimates used in the analysis from the best available source (RCT = best, expert opinion = worst)? | 8 | x |  |
| 4. | If estimates came from a subgroup analysis, were the groups pre-specified at the beginning of the study? | 1 | x |  |
| 5. | Was uncertainty handled by (1) statistical analysis to address random events, (2) sensitivity analysis to cover a range of assumptions? | 9 | x |  |
| 6. | Was incremental analysis performed between alternatives for resources and costs? | 6 | x |  |
| 7. | Was the methodology for data extraction (including the value of health states and other benefits) stated? | 5 | x |  |
| 8. | Did the analytic horizon allow time for all relevant and important outcomes? Were benefits and costs that went beyond 1 year discounted (3% - 5%) and justification given for the discount rate? | 7 | x |  |
| 9. | Was the measurement of costs appropriate and the methodology for the estimation of quantities and unit costs clearly described? | 8 | x |  |
| 10. | Was the primary outcomes measure(s) for the economic evaluation clearly stated and were the major short-term, long-term and negative outcomes included? | 6 |  | x |
| 11. | Were the health outcomes measures/scales valid and reliable? If previously tested valid and reliable measures were not available, was justification given for the measures/scales used? | 7 | x |  |
| 12. | Were the economic model (including structure), study methods and analysis, and the components of the numerator and denominator displayed in a clear, transparent manner? | 8 | x |  |
| 13. | Were the choice of economic model, main assumptions, and limitations of the study stated and justified? | 7 | x |  |
| 14. | Did the author(s) explicitly discuss the direction and magnitude of potential biases? | 6 | x |  |
| 15. | Were the conclusions/recommendations of the study justified and based on the study results? | 8 | x |  |
| 16. | Was there a statement disclosing the source of funding for the study? | 3 | x |  |
| Total score | | 100 | 94 |  |

**Quality of Health Economic Studies (QHES) instrument – Pashayan et al, 2018**

|  | Questions | Points available | Yes | No |
| --- | --- | --- | --- | --- |
| 1. | Was the study objective presented in a clear, specific, and measurable manner? | 7 | x |  |
| 2. | Were the perspective of the analysis (societal, third-party payer, etc.) and reasons for its selection stated? | 4 | x |  |
| 3. | Were variable estimates used in the analysis from the best available source (RCT = best, expert opinion = worst)? | 8 | x |  |
| 4. | If estimates came from a subgroup analysis, were the groups pre-specified at the beginning of the study? | 1 | x |  |
| 5. | Was uncertainty handled by (1) statistical analysis to address random events, (2) sensitivity analysis to cover a range of assumptions? | 9 | x |  |
| 6. | Was incremental analysis performed between alternatives for resources and costs? | 6 | x |  |
| 7. | Was the methodology for data extraction (including the value of health states and other benefits) stated? | 5 | x |  |
| 8. | Did the analytic horizon allow time for all relevant and important outcomes? Were benefits and costs that went beyond 1 year discounted (3% - 5%) and justification given for the discount rate? | 7 | x |  |
| 9. | Was the measurement of costs appropriate and the methodology for the estimation of quantities and unit costs clearly described? | 8 | x |  |
| 10. | Was the primary outcomes measure(s) for the economic evaluation clearly stated and were the major short-term, long-term and negative outcomes included? | 6 |  | x |
| 11. | Were the health outcomes measures/scales valid and reliable? If previously tested valid and reliable measures were not available, was justification given for the measures/scales used? | 7 | x |  |
| 12. | Were the economic model (including structure), study methods and analysis, and the components of the numerator and denominator displayed in a clear, transparent manner? | 8 | x |  |
| 13. | Were the choice of economic model, main assumptions, and limitations of the study stated and justified? | 7 | x |  |
| 14. | Did the author(s) explicitly discuss the direction and magnitude of potential biases? | 6 | x |  |
| 15. | Were the conclusions/recommendations of the study justified and based on the study results? | 8 | x |  |
| 16. | Was there a statement disclosing the source of funding for the study? | 3 | x |  |
| Total score | | 100 | 94 |  |
